# Study Protocol for the Intraoperative Complications Assessment and Reporting with Universal Standards (ICARUS) Global Cross-Specialty Surveys Among Surgeons, Anesthesiologists, Nurses, Interventional Cardiologists, and Interventional Radiologists

**DOI:** 10.1101/2023.08.08.23293789

**Authors:** Giovanni E. Cacciamani, Tamir Sholklapper, Michael B. Eppler, Aref Sayegh, Lorenzo Stornino Ramacciotti, Andre L. Abreu, Rene Sotelo, Mihir M. Desai, Inderbir S. Gill, ICARUS Global Surgical Collaboration

**Affiliations:** USC Institute of Urology and Catherine and Joseph Aresty Department of Urology, Keck School of Medicine, University of Southern California, Los Angeles, CA, USA; Artificial Intelligence Center at USC Urology, USC Institute of Urology, University of Southern California, Los Angeles, CA, USA; Norris Cancer Center, Keck School of Medicine, University of Southern California, Los Angeles, CA, USA; Department of Urology, Einstein Healthcare Network, Philadelphia, PA, USA; Department of Surgery, MedStar Good Samaritan Hospital, Baltimore, MD, USA

**Keywords:** anesthesiology, complications, ICARUS, intraoperative adverse events, intraoperative complications, surgery, surgical error

## Abstract

Every year, approximately 200 million surgeries are performed worldwide, and intraoperative adverse events (iAEs) have a significant impact on patients and surgeons. Despite their importance, the true scale of iAEs remains underestimated due to inadequate methods for assessment, collection, grading, and reporting. Various grading systems have been introduced over the past decade, but their adoption has been limited, leading to inconsistencies in reporting. Furthermore, a lack of standardized frameworks for defining, assessing, and collecting iAEs, coupled with litigation concerns, contributes to underreporting. Only half of surgery and anesthesiology journals provide guidance on reporting perioperative adverse events, and recommendations for reporting iAEs are notably lacking in surgical literature. To address these issues, the Intraoperative Complications Assessment and Reporting with Universal Standard (ICARUS) Global Surgical Collaboration was established in 2022. The initiative involves conducting global surveys and a Delphi consensus to understand the barriers for poor reporting of iAEs, validate shared criteria for reporting, define iAEs according to surgical procedures, evaluate the existing grading systems’ reliability, and identify strategies for enhancing the collection, reporting, and management of iAEs. A sample size of 2,398 respondents was calculated for the study, with invitations extended to 86,574 healthcare providers. This effort represents an essential step towards improved patient safety and the well-being of healthcare professionals in the surgical field.

## INTRODUCTION

Every year, approximately 200 million surgeries are performed worldwide[1, 2]. Intraoperative adverse events (iAEs) can occur, affecting patients’ perioperative outcomes and survival as well as the well-being of surgeons [3, 4]. Despite their significance, the true scale of iAEs remains underestimated due to inadequate methods for assessment, collection, grading, and reporting.

Given that these events “*do not stay in the OR*,” [5] a more accurate understanding of their prevalence could aid in developing strategies to prevent them and pathways for their management [3, 6, 7].

Over the past decade, different grading systems have been introduced to capture iAEs [8, 9] However, their adoption has been limited, and the lack of consensus across specialties necessitates external assessments [8]. Additionally, the absence of a clear, standardized definition of iAEs and their effects on postoperative courses has led to inconsistency in reporting. Nowadays, recommendations for reporting adverse events are lacking in surgical literature [4, 10]. Only half of the surgery and anesthesiology journals provide guidance on reporting perioperative adverse events, and less than 0.5% recommend how to report intraoperative adverse events [11]. Not reporting these events does not mean that they didn’t happen. It simply means that they were not reported, possibly due to lack of proper tools and standardized frameworks for defining, assessing, grading and collecting them [12].

Apart from the lack of standardized definitions, grading, and reporting criteria, the insufficient evaluation of these events might also be potentially associated with the effects they impose on surgeons’ well-being. Preliminary findings from the pioneering BISA study [13] showed that only a handful of surgeon’s report iAEs, largely out of litigation concerns and the absence of a robust reporting system. These results highlight the urgency for a global validation process to acknowledge surgeons as “second victims” and to implement support mechanisms in the aftermath of iAEs.

In 2022, the Intraoperative Complications Assessment and Reporting with Universal Standard Global Surgical Collaboration was established [10, 14] to address these issues and enhance patient safety. As part of this initiative, three global surveys and a Delphi consensus are released. These efforts aim to 1) understand the barriers for the poor reporting of iAEs and their impact on healthcare professionals, 2) validate common-shared criteria for improved iAEs reporting, 3) define iAEs according to surgical procedures and related anesthesiologic services, evaluate the inter-rater reliability of existing grading systems, and 5) identify strategies to enhance the collection, reporting, and management of iAEs.

## METHODS

### Selection and recruitment

The determination of the necessary sample size was made to reach a 95% confidence level and a 2% margin of error, as outlined before [15]. According to the World Healthcare Organization (WHO) Surgical Workforce Census, there were 1,853,842 surgeons and anesthesiologists around the world (https://apps.who.int/gho/data/view.main.HRSWF), thus a minimum of 2,398 respondents were calculated for the study. Corresponding authors who had published in the top-10 journals related to anesthesiology and surgery, intervention radiology and interventional cardiology between the years 2019 and 2021 were invited to participate through email. The identification of these journals and their rankings were done using the SCiMago Database (https://www.scimagojr.com). Specifically, these journals ware associated with one or more of the following specialties: Anesthesiology, Interventional Radiology, Interventional Cardiology, Nursing, Cardiothoracic Surgery, Colon and Rectal Surgery, General Surgery, Gynecologic Oncology, Gynecology and Obstetrics, Neurological Surgery, Ophthalmologic Surgery, Oral and Maxillofacial Surgery, Orthopaedic Surgery, Otorhinolaryngology, Plastic Surgery, Urology, Vascular Surgery. We utilized snowball sampling, encouraging respondents to share the survey with their colleagues and though social media distribution as previously done [16]. In total, 86,574 healthcare providers, consisting of 82,598 corresponding authors and 3,976 referees, were reached out to for participation in the surveys. This number does not include any additional participants from snowball sampling, or account for potential typos or defunct email addresses. To achieve a well-rounded representation encompassing diverse viewpoints, surgical specialties, and a wide spectrum of stakeholders, we will implement a purposeful oversampling strategy. This approach guarantees the inclusion of voices from different fields, locations, and demographic groups. By taking these steps, we intend to assemble a varied and inclusive set of participants, thus boosting the reliability, relevance, and validity of our research findings. Summary of type of surveys characteristics, objectives, rewards, and registration numbers is provided in *table 1*.

For each survey, baselines characteristics as detailed in *table 2* were collected.

Details about the aims, list of domains evaluated, and the type of analysis for each survey are reported separately as follows.

### Survey 1: Intraoperative Adverse Events Experience, Perception and Reporting

#### Aims

The purpose of this survey is three-fold. First (a), to better understand surgeons, anesthesiologists, interventional cardiologists, interventional radiologists, and nurses’ perceptions and experiences surrounding iAEs event reporting. Second (b), is to globally validate the utility of a recently developed ICARUS Global Surgical collaboration Criteria [14, 17] and to determine their applicability in various surgical specialties; lastly (c) to identify strategies and tools to improve the iAEs collection and reporting. Respondents were asked the consent to be contacted for follow-up studies (survey 2 and 3).

#### Survey areas, domains, and assessment

The survey is divided in the 3 main areas (a-c) to compile with the primary study objectives as reported in *table 1*.

a. *Intraoperative Adverse Event (iAE) Experience and Perception* Overall, the section seeks to gather comprehensive information about the participants’ interaction with iAEs, from their practical handling of these events to their emotional responses and beliefs about the implications of reporting such events. It also aims to understand the systemic support or challenges faced, as well as gather suggestions for improvements in the reporting process. Full list of questions is reported in the supplementary materials. Questions are shaped on a preliminary pilot study [10] and based on the Boston Intraoperative Adverse Events Surgeons’ Attitude survey [13] and on a previews review on the potential consequences of patient’s complications on surgeon wellbeing [18]. The questions in this section are designed to examine the participants’ experiences, practices, beliefs, and emotions surrounding intraoperative adverse events (iAEs). The domains surveyed in this section and their assessment is reported in *table 3*.
b. *Intraoperative adverse Events (iAE) Collection and Reporting* This section of the survey 1 aims to perform a worldwide cross-specialty assessment of the global applicability of a core set of criteria for reporting iAEs in clinical studies [10]. The focus of this assessment is to understand how universally these criteria can be applied by considering factors such as clarity, exhaustiveness, clinical usefulness, and quality assessment. The goal is to establish standardized guidelines that can be adopted universally, enhancing the consistency and quality of iAE reporting worldwide [14]. The reliability and consistency between different raters are evaluated to confirm the level of agreement. To calculate the percentage of agreement, Likert scale responses are divided into two categories: scores of 4 (useful) or 5 (very useful) represent agreement, while scores of 1 (not useful), 2 (less useful), or 3 (neutral) are taken as indicative of disagreement. The validity of the criteria is assessed in 5 domains with 5-point Likert scale responses: clarity, exhaustiveness, clinical utility, quality assessment and improvement utility, and research utility [19, 20]. Full list of questions is reported in the supplementary materials. The domains surveyed and their assessment in this section can be grouped as reported in *table 4*.
c. *Strategies/tools to improve the iAEs collection and reporting*. This section of survey 1 goal of this section is to assess and gather feedback on various methods and tools for the assessment, grading, and reporting of intraoperative adverse events (iAEs). This includes the collection of data, standardization of reporting, efficiency of tools, and potential additional resources. Domains and their assessment are reported in *table 5*.

#### Statistical analysis

Continuous and dichotomous variables are presented as median, mean (SD), and percentages when appropriate. Inter-rater reliability and the consistency of responses are assessed to establish the level of agreement. In the determination of the percentage of agreement, Likert responses are dichotomized, with scores of 4 (useful) or 5 (very useful) indicating agreement, and scores of 1 (not useful), 2 (less useful), or 3 (neutral) indicating disagreement. Screening for outliers is performed through the evaluation of absolute individual question agreement and the distribution of responses. The internal consistency is evaluated using Cronbach’s _α_ [21]. For purposes of global applicability of ICARUS Global Surgical collaboration criteria (domain b) as above reported), a minimum of 80% agreement and appropriate interrater consistency (Cronbach’s _α_ >0.5) is required in at least 3 of the 5 domains. At least 1 of the 3 (or more) domains to achieve the minimums must include clinical utility, quality assessment, and improvement utility, and research utility. Sub-group analysis is performed for each of the surveyed specialties.

#### Survey Distribution

Google™ Forms (https://docs.google.com/forms/) is the platform used to distribute, collect, and handle the study data. The method of snowball sampling is in place to engage additional respondents. Participants must meet the following criteria to be included in the study: they must understand and willingly agree to participate; they should be proficient in English or have fluency in English medical terminology; and they must have current or previous experience with procedures or surgeries, irrespective of the specific field or domain. Responders are offered to be included in the acknowledgments section of the publications using the data retrieved, to compile with the ICJME criteria for authorship (*Supplementary – invitation email*), Table 1.

#### Ethical considerations and dissemination

This study is approved by the institutional IRB (UP-21-00473) and registered to ClinicalTrials.gov (NCT04994392). The survey outcomes will be disseminated through peer-reviewed journals, conference presentations, workshops, and webinars. A copy of the ICARUS Global surgical collaboration checklist validated though the cross specialty global survey will be uploaded in the EQUATOR Network website.

### Survey 2: Intraoperative Adverse Events Definitions

#### Aim

In the last two decades, the field of surgical care has grappled with the inconsistency of definitions surrounding “surgical error” and adverse events (AEs) [22, 23]. Such heterogeneity has impacted the quality of reporting and interpretation of scientific findings, reflecting the need for standardized terminology[24, 25]. Despite efforts to standardize definitions, especially pertaining to i AEs, wide acceptance of these definitions has remained elusive[26]. Current definitions for surgical complications may vary in scope and interpretation across different contexts, including preoperative, intraoperative, and postoperative stages d[27-30]. In addition, the lack of common ground in defining iAEs may contribute to underreporting[31]. Given this backdrop, the main goal of the study is to develop a globally recognized, standardized definition of the iAEs through a Delphi consensus. This effort is not only aimed at harmonizing clinical practice and research but also at providing a comprehensive framework that encompasses various stages and aspects of surgical care, from anesthesia to postoperative events.

#### Survey areas, domains, and assessment

This international, cross-disciplinary consensus puts forward a foundational set of definitions for day-of-surgery adverse events (DOS-AEs), encompassing preoperative AEs, intraoperative AEs, and immediate postoperative stages. This set takes into consideration the timing of AE occurrence, type of surgical/interventional procedure, the specific type of AE, and the quality of AE. It includes events related to surgical or interventional procedures, anesthesiology, and nursing, and it covers both harmful and potentially harmful occurrences. In the subsequent sections, we will provide Table 6 details domains and assessment regarding each of these mentioned aspects.

#### Modality

The Delphi survey will consist of several phases, during which the panelists will evaluate and anonymously choose to agree, disagree, or propose changes to the definition essential elements. This process will be carried out through a maximum of three rounds. Following each round, participants will be given collective feedback from the prior round, assisting in the alignment of individual opinions and the formation of a consensus within the group. Participants will assess the relevance and quality of essential elements in each in each definition for inclusion using a 1 to 5 Likert scale. A 5-point Likert scale is selected to measure consensus on each important element in the definition, with references [32, 33]. The numerical scores are defined as follows: 1: Strong agreement 2: Mild agreement 3: Indecision 4: Mild disagreement 5: Strong disagreement. In addition to utilizing the Likert scale, each question will provide a free-text space for participants to offer suggestions for improving the proposed definition or adding extra components. Participants will also have the option to select ‘unable/unwilling to answer’ for any of the questions.

#### Analysis

The agreement with definitions is being assessed on a 1 to 5-point Likert scale, and respondents who disagree, rating 1, 2, or 3, are asked to comment. Responses are analyzed, and agreement is considered achieved when 80% or more of the participants rate the criteria as 4 (agree) or 5 (strongly agree) on the Likert scale. Comments from those who disagree with the definitions are being reviewed by the study authors, and these comments are used to modify the definitions for the next round of the survey. Baseline characteristics are summarized by count and percent, and percent agreement is calculated as the proportion of those endorsing the definition with a 4 (agree) or 5 (strongly agree).

#### Survey Distribution

RedCap® is the platform used to distribute, collect, and handle the study data. The method of snowball sampling is in place to engage additional respondents. Participants must meet the following criteria to be included in the study: they must understand and willingly agree to participate; they should be proficient in English or have fluency in English medical terminology; and they must have current or previous experience with procedures or surgeries, irrespective of the specific field or domain. Responders who completed all the Delphi survey rounds, are offered collaborative authorship [34] of the publications using the data retrieved under the name *ICARUS Global Surgical Collaboration Research Group*, to compile with the ICJME criteria for authorship (*Supplementary – invitation email*), Table 1.

#### Ethical considerations and dissemination

This study is approved by the institutional IRB (UP-21-00473) and registered to ClinicalTrials.gov (NCT04994392). The survey outcomes will be disseminated through peer-reviewed journals, conference presentations, workshops, and webinars.

### Survey 3: Intraoperative Adverse Events Grading inter-rater reliability

#### Aims

Although there are various intraoperative grading and classification systems [8], informally referred to as: EAUiaiC, iAE severity classification scheme, Modified Satava, EAES Grading system, and ClassIntra® (previously known as CLASSIC), the documentation of these iAEs is still extremely rare. Additionally, while postoperative adverse events are commonly reported, only a small portion of surgical literature addresses intraoperative complications as noteworthy outcomes [9]. The chief goal of survey 3 is to assess the uniformity and inter-rater reliability of the 5 iAE grading systems regarding the distribution of responses by quantity and percentage. Results of this survey will be instrumental for understanding the external cross-specialty variability in grading these events using the existing iAEs grading systems.

#### Scenarios selection and assessment

Each of the iAEs grading systems that has been developed currently exhibits inter-rater reliability, assessed through carefully defined surgical and anesthesiological scenarios. To maintain consistency, and with the aim of contrasting the overall inter-rater reliability with the performance specific to iAEs, we compile all the scenarios from the various iAEs grading systems papers (total 68) into an Excel spreadsheet. Subsequently, utilizing Excel’s random sequence generation function, we produce a randomized selection of 10 distinct scenarios.

These selected scenarios are then independently assessed and graded employing all five of the existing iAEs grading systems. We invited the respondents to elucidate their comprehension of the iAEs scenario, with the intention of assessing both their understanding of the scenario itself and the consistency between this understanding and the potential heterogeneity in the inter-rater reliability associated with the utilization of the iAEs grading systems. The questions are shaped to compile with commonly shared domains utilized in each iAEs grading systems. The domains and corresponding assessments are systematically detailed in Table 7. Details of the questions are reported in supplementary material.

#### Statistical Analysis

The primary data analysis requires computations to examine the uniformity and inter-rater concordance for each grading system, as detailed below:

- Distribution of responses by quantity and percentage
- Consistency and inter-rater reliability assessment of the 5 iAE grading systems concerning the percentage agreement of grade.
- Consistency and inter-rater reliability evaluation of the 5 iAE grading systems employing Cohen’s K
- Consistency and inter-rater reliability examination of the 5 iAE grading systems utilizing the Intra-class correlation (ICC) with two-way, random effects to gauge the uniformity of grades.
- Comparison between inter-raters’ reliability in grading same scenarios will be performed
- Comparison between those respondents who already utilizes one of the grading systems vs. those who don’t’ will be performed.

#### Survey Distribution

RedCap ® is the platform used to distribute, collect, and handle the study data. The method of snowball sampling is in place to engage additional respondents. Participants must meet the following criteria to be included in the study: they must understand and willingly agree to participate; they should be proficient in English or have fluency in English medical terminology; and they must have current or previous experience with procedures or surgeries, irrespective of the specific field or domain. Responders are offered collaborative authorship [34] of the publications using the data retrieved under the name *ICARUS Global Surgical Collaboration Research Group*, to compile with the ICJME criteria for authorship (*Supplementary – invitation email*), Table 1.

#### Ethical considerations and dissemination

This study is approved by the institutional IRB (UP-21-01010) and registered to ClinicalTrials.gov (NCT05270603). The survey outcomes will be disseminated through peer-reviewed journals, conference presentations, workshops, and webinars.

## RESULTS REPORTING

The results of the surveys 1,2 and 3 will be reported separately in different publications and the reporting of the survey follow the Checklist for Reporting Results of Internet E-Surveys (CHERRIES)[35] and the American Association for public opinion Research (AAPOR) Survey Disclosure Checklist. The studies are formulated to address all applicable disclosure elements set forth by the American Association for Public Opinion Research Transparency Initiative.

## DISCUSSION

Intraoperative adverse events are poorly reported, and their impact on both patients and surgeons is often overlooked. The goal of the ICARUS Global Surgical Collaboration project is to create an ecosystem that enhances the assessment, grading, and reporting of iAEs. This improvement aims to evaluate their impact on patients and providers, and to establish frameworks that assist surgeons in handling these effects. The project also focuses on improving patient care by implementing standardized pathways to prevent iAEs and, if they occur, to manage and follow up on them.

In the present protocol study, we delineate the objectives, scope, and methodology for a series of three global surveys. These surveys are designed to investigate the underlying causes of the inadequate reporting of specific events and to develop universally accepted definitions and criteria to bolster the collection, assessment, and reporting process. By incorporating feedback from all healthcare providers, we aim to identify effective strategies to enhance current practices. The findings from these global surveys will be instrumental in formulating widely accepted guidelines, thereby improving the assessment of these events. Consequently, the insights gained will facilitate the creation of structured frameworks, leading to the advancement of patient care.

## Data Availability

All data produced in the present study are available upon reasonable request to the authors

