## Supplementary material for "Study Protocol for the Intraoperative Complications Assessment and Reporting with Universal Standards (ICARUS) Global Cross-Specialty Surveys Among Surgeons, Anesthesiologists, Nurses, Interventional Cardiologists, and Interventional Radiologists": Tables 1-7

**Table 1.** Summary of surveys objectives, rewards, and registration number. *Details of primary and secondary objectives are reported on ClinicalTrials.gov and in the supplementary materials. **Providers must complete all the rounds of the Delphi Consensus to be included in the authorship.

| **Survey** | **Objective(s)*** | **Reward** | **IRB Number** | **Clinicaltrial.gov Number** |
| --- | --- | --- | --- | --- |
| **Survey 1** | To understand the impact of iAEs on provider wellbeing. | Acknowledgement in the papers | UP-21-00473 | NCT04994392 |
|  | To understand the reasons for the poor reporting of iAEs. |  |  |  |
|  | To perform a global, cross-specialty validation of the ICARUS Criteria. |  |  |  |
|  | To involve providers interested in participating in the Delphi Consensus Survey (Survey 2) |  |  |  |
|  | Developing an ecosystem for effective data collection related to iAEs |  |  |  |
| **Survey 2** | Performing a comprehensive, cross-specialty definition of Day-of-surgery AEs with a modified Delphi framework | Collaborative authorship in the papers** |  |  |
| **Survey 3** | Evaluating the inter-rater reliability of iAEs grading systems. | Collaborative authorship in the papers | UP-21-01010 | NCT05270603 |
|  | Identifying common patterns to use for proposing a new iAEs grading system |  |  |  |

**Table 2** Demographic domains and assessment

| **Domains** | **Assessment** |
| --- | --- |
| **Age** | Assess the respondent's age. |
| **Country** | Determine the country in which the respondent practices. |
| **Practice years** | Identify approximately how many years the respondent has been practicing. |
| **Role** | Determine the current job title or role of the respondent. |
| **Specialty** | Identify which description most closely aligns with the respondent's specialty. |
| **Surgical Approach** | Determine the surgical approach the respondent uses (or participates in) in their practice. |
| **Practice Setting** | Identify the description that best matches the respondent's practice setting. |
| **Annual Surgical Volume** | Assess approximately how many procedures or surgeries the respondent performs (or participates in) annually. |
| **Main Area of Interest** | Determine the main area of interest and daily practice of the respondent. |

**Table 3.** Intraoperative Adverse Event (iAE) Experience and Perception; domains and assessment

| **Domain** | **Assessment** |
| --- | --- |
| **1. Experiences and Practices** | Frequency of witnessing iAEs in the past 12 months; Responses to iAEs; Importance of communication during or after iAEs; Debriefing and sharing practices with procedural teams, patients, and colleagues; Reporting and collecting iAEs in daily practice; Importance of regular collection of iAEs; Systems for grading iAEs. |
| **2. Practical Considerations** | Observations of iAEs; Standardized systems for assessing, reporting, and grading iAEs; Confidence in assessing and reporting iAEs; Mechanisms and support for addressing identified iAEs; Additional practical considerations impacting ability to report iAEs. |
| **3. Emotional Considerations** | Concerns about negative impacts on emotional well-being, self-confidence, and job performance; Emotional repercussions after iAEs; Additional emotional considerations related to iAE reporting. |
| **4. Perceived Benefits** | Beliefs about the clinical utility, quality assessment, enhancement of safety culture, patient safety, and educational value of reporting iAEs; Additional benefits to reporting iAEs. |
| **5. Perceived Consequences**  **to Clinical Practice** | Potential negative impacts on risk-taking and quality of surgical practice; Additional potential consequences affecting the decision to report iAEs. |
| **6. Improvement Suggestions** | Indicators for improving or increasing iAE reporting, both in personal practice and globally; Additional comments, suggestions, questions, and concerns related to iAE reporting. |

**Table 4.** Intraoperative adverse Events (iAE) Collection and Reporting. Domains and assessment accordingly to the ICARUS Global Surgical Collaboration Criteria [10]

| **Domain** | **Assessment** |
| --- | --- |
| **Reporting Intraoperative Adverse Events (IAEs)** | Focuses on the necessity to include IAEs as an essential outcome in perioperative study reports. |
| **Definition and Reference for IAEs** | IAEs and the definition of each specific IAE must be provided or referenced. |
| **Classification Systems for IAEs** | Each IAE should be reported using one of the proposed iAEs classification systems |
| **Reporting IAEs by Grade** | Focuses on reporting each IAE separately by grade. |
| **Anesthesiological and Surgical Complications** | Emphasizes the separate reporting of anesthesiological and surgical complications. |
| **Number of IAEs and Number of Patients** | Specifies the need to report the number of IAEs and the number of patients experiencing them separately. |
| **Conditions Associated with IAEs** | Underlines the importance of reporting conditions associated with IAEs when appropriate, following the standard criteria. |
| **IAE Conversion Reporting** | Discusses the necessity to report IAEs that require a conversion and the corresponding action taken, following the standard criteria. |
| **Surgical Step Associated with IAEs** | Calls for the reporting of the surgical step that was associated with or affected by the IAEs, in line with the usual criteria. |
| **Timing of IAEs Assessment** | Requires the timing of the IAEs assessment to be reported. |
| **Management of IAEs** | Focuses on the need to report the management of IAEs. |
| **Sequelae of IAEs in Postoperative Course** | Requires reporting of the sequelae of IAEs in the postoperative course, with compatibility with existing classification systems. |

**Table 5.** Strategies/tools to improve the iAEs collection and reporting. Domains and assessment

| **Domain** | **Assessment** |
| --- | --- |
| **Assessment via Patient Form** | Measures the efficacy of using patient forms in capturing data for the assessment, grading, and reporting of intraoperative adverse events. This could include self-reported data or questionnaires filled out by OR staff. |
| **Utilization of Online Grade Calculator/Converter** | Evaluates the usefulness of an online tool that calculates or converts data in order to grade and report iAEs. This could streamline data processing and standardize grading. |
| **Automated Data Recording for iAEs** | Examines the potential benefit of employing automated systems in recording data related to iAEs, which could minimize human error and enhance efficiency. |
| **Post-operative Time-out or Checklist Application** | Investigates the importance of using a checklist or time-out procedures after surgery to ensure proper data collection, and thereby assists in the accurate assessment, grading, and reporting of iAEs. |
| **Additional Resources for Data Collection** | Solicits suggestions for additional tools or resources that might be helpful in the data collection process for assessing, grading, and reporting iAEs. |
| **Scientific Publication & Criteria Checklist** | Assesses the value of having a standard criteria checklist for scientific publication of iAEs, enhancing uniformity and quality in published research. |
| **Importance of Guideline Recommendations by Academic Journals** | Evaluates how crucial it is for academic journals to offer standardized guidelines and recommendations for reporting iAEs, which can contribute to the credibility and consistency across the scientific community. |
| **Additional Resources for Scientific Publication** | Asks for extra resources or tools that might be beneficial in the process of scientifically publishing assessments, grades, and reports of iAEs, aiming to improve overall quality and consistency in academic work. |

**Table 6.** Day of surgery adverse events definitions. Domains and assessment

| **Domain** | **Assessment** |
| --- | --- |
| **Timing** | Occurring of AEs during the preoperative, intraoperative of immediately postoperative period (24 h after surgery), specifically during anesthesia and/or the surgical/interventional procedure time is assessed.  Differentiation between Anesthesia and Surgical times is tested with definitions as mentioned below. *Anesthesia Time* is a continuous time from the start of anesthesia to the end of an anesthesia service (as defined by 2019 ASA RVG).  Surgical/Interventional Procedure Time is defined as the time frame from when the initial incision was made for the principal procedure to skin closure. In the case of endoscopic maneuvers which do not require any skin incision (i.e.: cystoscopy, colonoscopy, etc.) the surgical procedure time is defined as the time frame from when the endoscopic tool is initially inserted for the principal procedure through the natural or pre-existing orifice to its withdrawal.) |
| **Nature** | The AEs may be unintended, unplanned, and possibly anticipable or preventable. Differentiation between "event" and its "effect" is tested |
| **Type of Procedures Involved** | Encompasses AEs occurring during anesthesia, surgical, or interventional procedures. For iAEs, the type of surgery/intervention is also considered as Surgical Procedures Under General Anesthesia; Surgical Procedures Under Sedation; Surgical Procedures Under Local Anesthesia; Surgical Procedures without Anesthesia. |
| **Potential Harm** | AEs is potentially harmful to the patient, indicating a focus on patient safety and well-being. |
| **Recognition** | AEs may be recognized either during or after the period of interest (as reported in the domain "timing"), signifying a domain related to the detection and awareness of the event. |

**Table 7**. General questions for iAEs comprehension evaluation. Domains and assessment

| **Domain** | **Assessment** |
| --- | --- |
| Grading system usage | Which intraoperative adverse event grading system(s) do you use, if used? |
| Patient outcomes | Was the iAE associated with death of the patient? |
|  | Was the iAE immediately life-threatening? |
|  | Were there significant consequences to the patient due to the iAE? |
| Surgical errors and mishaps | Was the incorrect site, side, or surgical approach used without consent? |
|  | Was the intraoperative injury missed, necessitating re-operation within 7 days of index procedure? |
| Procedural alterations and deviations | Were there any changes in the ideal intraoperative course related to iAE? |
|  | Was there an unanticipated conversion of approach or significant change in planned procedure due to iAE? |
|  | Was planned procedure aborted or incomplete due to iAE? |
|  | Unplanned stoma as a result of iAE? |
|  | Unplanned tissue or organ removal as a result of iAE? |
|  | Was any surgical repair, medical treatment, or other intervention required? |
|  | Was blood loss appreciably over normal range for procedure? |
|  | Were 2 or more units of blood products required to manage iAE? |
| Post-operative outcomes and care | Was there a change in post-operative care due to the iAE? |
|  | Did the iAE or its management necessitate intensive care admission? |
