## Supplementary materials for "Study Protocol for the Intraoperative Complications Assessment and Reporting with Universal Standards (ICARUS) Global Cross-Specialty Surveys Among Surgeons, Anesthesiologists, Nurses, Interventional Cardiologists, and Interventional Radiologists"

ICARUS Project Collaboration - Cross-specialty Global Validation Assessment

Dear Colleague,

I hope this email finds you well.

The lack of **intraoperative adverse event** assessment and reporting has been a significant gap in our literature for many years. As such, our team has been working to propose a list of standardized criteria to improve the **Intraoperative Complication Assessment and Reporting with Universal Standards (ICARUS).**

The goal of the **ICARUS project** - recently **listed in EQUATOR Network** as “Guidelines under Development” and in **ClinicalTrials.gov (NCT049943920)**- is to provide guidance for reporting intraoperative complications in research papers about surgical procedures in general. We hope these resources will improve our understanding of the nature and frequency of iAEs and increase our ability to counsel patients regarding surgical procedures.

In order to perform a Cross-Specialty Global Validation Assessment, **we invite you to please complete the survey on intraoperative adverse event (iAE) reporting**. It should take **approximately 10 minutes**, and it is intended for surgeons, anesthesiologists, and nurses with any level of experience in the operative setting.

ICARUS Questionnaire Link

Upon completing **all** questions, optional **identification details** will be collected, and you **will be automatically listed in acknowledgments of all the publications.**

Formal consent for survey participation will be assumed upon final submission.

Your participation is greatly appreciated.

Thank you!

Giovanni E. Cacciamani, MD
On behalf of the *ICARUS Project Steering Committee*

EQUATOR Network Submission - ClinicalTrials.gov (NCT049943920)


-----------------------------------------------------------------------------

**Giovanni E. Cacciamani, MSc, MD**
*Assistant Professor of Research Urology*
*Assistant Professor of Research Radiology*
*Co-Director of the AI Center of Urology*
*Vice-Chair of Research Council (Residents/Fellows)*
Institute of Urology, Keck School of Medicine
University of Southern California (USC)
Los Angeles, CA, USA

--------------------------------------------------------------------------------

ORCID: https://orcid.org/0000-0002-8892-5539
PubMed Library: https://www.ncbi.nlm.nih.gov/myncbi/giovanni%20enrico.cacciamani.1/bibliography/public/
Keck School of Medicine: https://keck.usc.edu/faculty-search/giovanni-cacciamani/
Twitter: https://twitter.com/Cacciamani_MD @Cacciamani_MD

US phone number +1 626 491 1531
IT phone number +39 340 24 91 750

### Surgeon, anesthesiologist, interventional radiologist, interventional cardiologist, and nurse experience with intraoperative adverse event (iAE) reporting

You have been selected to participate to the survey according to strict inclusion criteria [ClinicalTrials.gov (NCT049943920)]. Please do not share this questionnaire or part of it.

---

You are invited to participate in a research study. Your participation is voluntary. This document explains information about this study. You should ask questions about anything that is unclear to you.

In total, the survey should take no more than 10 minutes.

PURPOSE
The purpose of this study is two-fold. First, we hope to better understand surgeons, anesthesiologists, interventional cardiologists, interventional radiologists, and nurses' perceptions and experiences surrounding intraoperative adverse event reporting. Second, we hope to globally validate the utility of a recently developed intraoperative adverse event reporting guidelines, known as the Intraoperative Complication Assessment and Reporting with Universal Standards (ICARUS) Guidelines. We also hope to determine the applicability of the ICARUS guidelines in various surgical specialties. You are invited as a possible participant because of your surgical or procedural experience. The study is registered in ClinicalTrials.gov (NCT049943920)

This questionnaire is divided into seven parts as following

PART 1 - Demographics and expertise
PART 2 - Intraoperative Adverse Event (iAE) Definitions
PART 3 - Intraoperative Adverse Event (iAE) Experience
PART 4 - Intraoperative Adverse Event (iAE) Perception
PART 5 - How to improve Intraoperative Adverse Event (iAE) Reporting
PART 6 - ICARUS Reporting Guidelines Global Assessment
PART 7 - Resources and tools for Grading and Reporting the iAEs

PARTICIPANT INVOLVEMENT
Participants will be asked to answer a series of questions regarding their surgical or procedure practice and experience related to intraoperative adverse event reporting. Participants will also be asked to read and then evaluate a set of 13 intraoperative adverse event reporting criteria. There are no additional requirements for participation in this study beyond the completion of the survey.

PAYMENT/COMPENSATION FOR PARTICIPATION
Participation in this study is voluntary and there is no compensation for participation in this study.

ALTERNATIVES TO PARTICIPATION
You have the option to not participate in this study.

CONFIDENTIALITY
The members of the research team and the University of Southern California Institutional Review Board (IRB) may access the data. The IRB reviews and monitors research studies to protect the rights and welfare of research subjects.

Data collected in the survey will remain anonymous. No effort will be made to identify or contact participants.

If any personally identifiable information is found in the survey, it will immediately be removed by the study personnel and will not be stored.

Some participants may choose to provide their information for purposes of public acknowledgment of participation as part of a future study manuscript. In order to protect the anonymity of survey responses, this information will be collected separately from survey data.

INVESTIGATOR CONTACT INFORMATION
If you have any questions about this study, please contact the primary investigator Giovanni E. Cacciamani, MD by

IRB CONTACT INFORMATION
If you have any questions about your rights as a research participant, please contact the University of Southern California Institutional Review Board at (323) 442-0114 or.

* Indicates required question

Please note that all survey responses will be anonymized. If you are interested in having your name include in the acknowledgment of a future publication related to the ICARUS Survey, space will be provided at the completion of the survey.

Clicking "next" indicates that you have read and understood the above information and agree to participate i this study:

PART 1 - Demographics and expertise

1. What is your age? *

2. In which country do you practice? *

*Mark only one oval.*

- Afghanistan
- Akrotiri
- Albania
- Algeria
- American Samoa
- Andorra
- Angola
- Anguilla
- Antarctica
- Antigua and Barbuda
- Argentina
- Armenia
- Aruba
- Ashmore and Cartier Islands
- Australia
- Austria
- Azerbaijan
- Bahamas, The
- Bahrain
- Bangladesh
- Barbados
- Bassas da India
- Belarus
- Belgium
- Belize
- Benin
- Bermuda
- Bhutan
- Bolivia
- Bosnia and Herzegovina
- Botswana
- Bouvet Island

- Brazil
- British Indian Ocean Territory
- British Virgin Islands
- Brunei
- Bulgaria
- Burkina Faso
- Burma
- Burundi
- Cambodia
- Cameroon
- Canada
- Cape Verde
- Cayman Islands
- Central African Republic
- Chad
- Chile
- China
- Christmas Island
- Clipperton Island
- Cocos (Keeling) Islands
- Colombia
- Comoros
- Congo, Democratic Republic of the
- Congo, Republic of the
- Cook Islands
- Coral Sea Islands
- Costa Rica
- Cote d'Ivoire
- Croatia
- Cuba
- Cyprus
- Czech Republic

- Denmark
- Dhekelia
- Djibouti
- Dominica
- Dominican Republic
- Ecuador
- Egypt
- El Salvador
- Equatorial Guinea
- Eritrea
- Estonia
- Ethiopia
- Europa Island
- Falkland Islands (Islas Malvinas)
- Faroe Islands
- Fiji
- Finland
- France
- French Guiana
- French Polynesia
- French Southern and Antarctic Lands
- Gabon
- Gambia, The
- Gaza Strip
- Georgia
- Germany
- Ghana
- Gibraltar
- Glorioso Islands
- Greece
- Greenland
- Grenada

- Guadeloupe
- Guam
- Guatemala
- Guernsey
- Guinea
- Guinea-Bissau
- Guyana
- Haiti
- Heard Island and McDonald Islands
- Holy See (Vatican City)
- Honduras
- Hong Kong
- Hungary
- Iceland
- India
- Indonesia
- Iran
- Iraq
- Ireland
- Isle of Man
- Israel
- Italy
- Jamaica
- Jan Mayen
- Japan
- Jersey
- Jordan
- Juan de Nova Island
- Kazakhstan
- Kenya
- Kiribati
- Korea, North

- Korea, South
- Kuwait
- Kyrgyzstan
- Laos
- Latvia
- Lebanon
- Lesotho
- Liberia
- Libya
- Liechtenstein
- Lithuania
- Luxembourg
- Macau
- Macedonia
- Madagascar
- Malawi
- Malaysia
- Maldives
- Mali
- Malta
- Marshall Islands
- Martinique
- Mauritania
- Mauritius
- Mayotte
- Mexico
- Micronesia, Federated States of
- Moldova
- Monaco
- Mongolia
- Montenegro
- Montserrat

- Morocco
- Mozambique
- Namibia
- Nauru
- Navassa Island
- Nepal
- Netherlands
- Netherlands Antilles
- New Caledonia
- New Zealand
- Nicaragua
- Niger
- Nigeria
- Niue
- Norfolk Island
- Northern Mariana Islands
- Norway
- Oman
- Pakistan
- Palau
- Panama
- Papua New Guinea
- Paracel Islands
- Paraguay
- Peru
- Philippines
- Pitcairn Islands
- Poland
- Portugal
- Puerto Rico
- Qatar
- Reunion

- Romania
- Russia
- Rwanda
- Saint Helena
- Saint Kitts and Nevis
- Saint Lucia
- Saint Pierre and Miquelon
- Saint Vincent and the Grenadines
- Samoa
- San Marino
- Sao Tome and Principe
- Saudi Arabia
- Senegal
- Serbia
- Seychelles
- Sierra Leone
- Singapore
- Slovakia
- Slovenia
- Solomon Islands
- Somalia
- South Africa
- South Georgia and the South Sandwich Islands
- Spain
- Spratly Islands
- Sri Lanka
- Sudan
- Suriname
- Svalbard
- Swaziland
- Sweden
- Switzerland

- Syria
- Taiwan
- Tajikistan
- Tanzania
- Thailand
- Timor-Leste
- Togo
- Tokelau
- Tonga
- Trinidad and Tobago
- Tromelin Island
- Tunisia
- Turkey
- Turkmenistan
- Turks and Caicos Islands
- Tuvalu
- Uganda
- Ukraine
- United Arab Emirates
- United Kingdom
- United States
- Uruguay
- Uzbekistan
- Vanuatu
- Venezuela
- Vietnam
- Virgin Islands
- Wake Island
- Wallis and Futuna
- West Bank
- Western Sahara
- Yemen

- Zambia
- Zimbabwe
- Other

3. Approximately how many years have you been practicing? *

Including residency and other formal training

*Mark only one oval.*

- < 5
- 5 - 9
- 10 - 14
- 15 - 19
- 20 - 24
- > 25
- N/A

4. What is your current job title or role? *

*Mark only one oval.*

- Medical Student
- Resident
- Clinical Fellow
- Clinical instructor
- Faculty physician
- Consultant
- Nurse
- Researcher
- Other: ______________________

5. Which of the following most closely describes your specialty? *

*Mark only one oval.*

- Anesthesiology *Skip to question 10*
- Cardiothoracic surgery *Skip to question 10*
- Colon and rectal surgery *Skip to question 10*
- General surgery *Skip to question 10*
- Gynecologic oncology *Skip to question 10*
- Gynecology and obstetrics *Skip to question 10*
- Neurological surgery *Skip to question 10*
- Ophthalmologic surgery *Skip to question 10*
- Oral and maxillofacial surgery *Skip to question 10*
- Orthopaedic surgery *Skip to question 10*
- Otorhinolaryngology *Skip to question 10*
- Pediatric surgery *Skip to question 10*
- Plastic surgery *Skip to question 10*
- Surgical oncology *Skip to question 10*
- Transplant surgery *Skip to question 10*
- Trauma surgery *Skip to question 10*
- Urology
- Vascular surgery *Skip to question 10*
- Interventional Radiology *Skip to question 10*
- Interventional Cardiologist *Skip to question 10*
- Other: ______________________

6. What surgical approach do you use (or participate in) in your practice? *

*Check all that apply.*

- [ ] Open
- [ ] Laparoscopic
- [ ] Robotic
- [ ] Endoscopic
- [ ] Imaging guided
- [ ] External (e.g. dermatologic)
- [ ] Other: ____________________

7. Which best describes your practice setting? *

*Mark only one oval.*

- ( ) Academic
- ( ) Community based
- ( ) Hybrid community and academic
- ( ) Military
- ( ) Other: ____________________

8. Approximately how many procedures or surgeries do you perform (or participate in) annually? *

____________________

#### Urologic Sub-specialty

9. Which is your main area of interest and daily practice (select all that apply) *

*Check all that apply.*

- [ ] General urologic surgery
- [ ] Urologic oncology and urologic oncological surgery
- [ ] Endourology and endourologic surgery
- [ ] Urogynecology and urogynecologic surgery
- [ ] Reconstructive urologic surgery (a form of reconstructive surgery)
- [ ] Pediatric urology and pediatric urologic surgery
- [ ] Transplant urology
- [ ] Trauma urology surgery
- [ ] Functional urology (voiding dysfunction, paruresis, neurourology)
- [ ] Surgical andrology and sexual medicine
- [ ] Gender affirmation surgery
- [ ] Imaging Guided Diagnostic Surgery (Biopsy)
- [ ] Focal Therapy
- [ ] Other: ______________________

#### PART 2 - Intraoperative Adverse Event (iAE) Definitions

Using the scale, please indicate your agreement with the definition of iAEs as it relates to specific circumstances

10. GENERALLY, an Intraoperative Adverse Event (iAE) is "any unanticipated or unplanned incident, potentially harmful for the patient, that occurs during the OR time (regardless of the type of anesthesia).

*Mark only one oval per row.*

| | 5 = Strongly agree | 4 | 3 = Neither agree nor disagree | 2 | 1 = Strongly disagree |
|---|---|---|---|---|---|
| **Agreement with definition** | ◯ | ◯ | ◯ | ◯ | ◯ |

11. Comments on general definition (Optional)

12. In cases WITHOUT ANESTHESIA, an Intraoperative Adverse Event (iAE) is "any unanticipated or unplanned incident, potentially harmful for the patient, that occurs during the surgical procedure time" (Note: surgical procedure time is defined as the time the initial incision was made for the principal procedure to skin closure. In the case of endoscopic maneuvers which do not require any skin incisior (ie: cystoscopy, colonoscopy, etc) the surgical procedure time is defined as the time the initial endoscopic tool is inserted for the principal procedure through the natural or pre-existing orifice to its withdraw.)

*Mark only one oval per row.*

| | 5 = Strongly agree | 4 | 3 = Neither agree nor disagree | 2 | 1 = Strongly disagree |
|---|---|---|---|---|---|
| **Agreement with definition** | ◯ | ◯ | ◯ | ◯ | ◯ |

13. Comments on iAE without anesthesia definition (Optional)

14. In cases under LOCAL ANESTHESIA, an Intraoperative Adverse Event (iAE) is "any unanticipated or unplanned incident,potentially harmful for the patient, that occurs during the surgical procedure time" (Note: in this case, surgical procedure time is defined as the time from injection of local anesthetic to completion of the primary procedure)

*Mark only one oval per row.*

| | 5 = Strongly agree | 4 | 3 = Neither agree nor disagree | 2 | 1 = Strongly disagree |
|---|---|---|---|---|---|
| **Agreement with definition** | ◯ | ◯ | ◯ | ◯ | ◯ |

15. Comments on iAE under local anesthesia definition (Optional)

_______________________________________________

_______________________________________________

_______________________________________________

_______________________________________________

_______________________________________________

16. In cases under SEDATION, an Intraoperative Adverse Event (iAE) is "any unanticipated or unplanned incident, potentially harmful for the patient, that occurs during the ANESTHESIA TIME - which includes the Surgery Time". (Note: Anesthesia time is a continuous time period from the start of anesthesia to the end of an anesthesia service)

*Mark only one oval per row.*

| | 5 = Strongly agree | 4 | 3 = Neither agree nor disagree | 2 | 1 = Strongly disagree |
|---|---|---|---|---|---|
| **Agreement with definition** | ◯ | ◯ | ◯ | ◯ | ◯ |

17. Comments on iAE under SEDATION definition (Optional)

18. In cases under GENERAL ANESTHESIA, an Intraoperative Adverse Event (iAE) is "any unanticipated or unplanned incident, potentially harmful for the patient, that occurs during the ANESTHESIA TIME which includes the Surgery Time". (Note: Anesthesia time is a continuous time period from the start of anesthesia to the end of an anesthesia service)

*Mark only one oval per row.*

| | 5 = Strongly agree | 4 | 3 = Neither agree nor disagree | 2 | 1 = Strongly disagree |
|---|---|---|---|---|---|
| **Agreement with definition** | ◯ | ◯ | ◯ | ◯ | ◯ |

19. Comments on iAE under general anesthesia definition (Optional)

20. Generally, a Perioperative Adverse event is "any immediate unanticipated or unplanned incident that occurs within 24h after the surgical procedure time or anesthesia time". (Note: definition of surgical procedure time and anesthesia time as reported above)

*Mark only one oval per row.*

| | 5 = Strongly agree | 4 | 3 = Neither agree nor disagree | 2 | 1 = Strongly disagree |
|---|---|---|---|---|---|
| **Agreement with definition** | ⬭ | ⬭ | ⬭ | ⬭ | ⬭ |

21. Comments on Perioperative Adverse Event definition (Optional)

PART 3 - Intraoperative Adverse Event (iAE) Experience

22. Approximately how many intraoperative adverse events have you witnessed in the past 12 months? *
Include events witnessed both personally and colleagues

23. What has been your response during these iAEs (select all that apply) *

*Check all that apply.*

- [ ] I have not observed any iAEs
- [ ] Attempted to address the iAE myself
- [ ] Communicated with the OR nursing staff regarding the iAE
- [ ] Communicated with the surgical team regarding the iAE
- [ ] Communicated with the anesthesia team regarding the iAE
- [ ] Other: ______________________

24. Do you believe communication about iAEs during or immediately after the procedure is important *

*Mark only one oval.*

- ( ) Yes
- ( ) No
- ( ) Other: ______________________

25. Do you de-brief with the procedural team to discuss iAEs after case completion? *

*Mark only one oval.*

- ( ) Yes, always
- ( ) Yes, only if time permits
- ( ) Yes, only if it is clinically relevant
- ( ) Never
- ( ) Other: ______________________

26. Do you share an observed iAE with your patients after the procedure? *

*Mark only one oval.*

- Yes, always
- Yes, only if time permits
- Yes, only if it is clinically relevant
- Never
- Other: ____________

27. Do you share your iAE experience with your colleagues who were not present during the procedure? *

*Mark only one oval.*

- Yes, always
- Yes, only if time permits
- Yes, only if it is clinically relevant
- Never
- Other: ____________

28. Where do you report intraoperative adverse events in your daily practice? *

*Check all that apply.*

- [ ] Operative Report
- [ ] Anesthesia Report
- [ ] Patient Charts
- [ ] A dedicated "Patient Form"
- [ ] Event reporting system
- [ ] Morbidity and mortality conferences
- [ ] Other: ____________

29. Do you collect/report intraoperative adverse events in your daily practice? *

*Mark only one oval.*

- ( ) Yes, always
- ( ) Yes, if it is clinically relevant
- ( ) No *Skip to question 32*

30. How important do you believe it is to regularly collect intraoperative adverse events? *

*Mark only one oval.*

| | 1 | 2 | 3 | 4 | 5 | |
|---|---|---|---|---|---|---|
| Not | ( ) | ( ) | ( ) | ( ) | ( ) | Extremely important |

#### Intraoperative Adverse Event (iAE) Grading

31. Which intraoperative adverse event grading system do you use?

*Check all that apply.*

- [ ] None, I document the event without grading
- [ ] EAUiaiC
- [ ] Modified Satava
- [ ] iAE Severity
- [ ] CLASSIC
- [ ] ClassIntra (Formerly CLASSIC)
- [ ] Clavien-Dindo
- [ ] Other: ____________________

#### PART 4 - Intraoperative Adverse Event (iAE) Perceptions

Using the scale, please indicate your agreement with the below statements as they related to iAE reporting.

32. Practical considerations *

*Mark only one oval per row.*

| | 5 = Strongly agree / Very much describes my practice | 4 | 3 = Neither agree nor disagree / this may or may not describe my practice, or my practice has too much variation | 2 | 1 = Strongly disagree / does not describe my practice | N/A |
|---|---|---|---|---|---|---|
| **I regularly observe intraprocedural complications** | ⬭ | ⬭ | ⬭ | ⬭ | ⬭ | ⬭ |
| **My practice has a standardized system for assessing, reporting and grading these complications** | ⬭ | ⬭ | ⬭ | ⬭ | ⬭ | ⬭ |
| **I feel confident in my ability to objectively assess and report these complications** | ⬭ | ⬭ | ⬭ | ⬭ | ⬭ | ⬭ |
| **My practice has appropriate mechanisms to act on any identified complications** | ⬭ | ⬭ | ⬭ | ⬭ | ⬭ | ⬭ |
| **My practice has the support necessary to report these complications** | ⬭ | ⬭ | ⬭ | ⬭ | ⬭ | ⬭ |

33. Please specify any additional practical considerations that either support or challenge your ability to report intraoperative adverse events:

If any of the above are Not Applicable, please specify below

34. Emotional considerations *

*Mark only one oval per row.*

| | 5 = Strongly agree / Very much describes my practice | 4 | 3 = Neither agree nor disagree / this may or may not describe my practice, or my practice has too much variation | 2 | 1 = Strongly disagree / does not describe my practice | N/A |
|---|---|---|---|---|---|---|
| **I am concerned that reporting these complications may negatively impact my emotional well-being** | ⬭ | ⬭ | ⬭ | ⬭ | ⬭ | ⬭ |
| **I am concerned that reporting these complications may negatively impact my self-confidence** | ⬭ | ⬭ | ⬭ | ⬭ | ⬭ | ⬭ |
| **I am concerned that reporting these complications may negative impact my ability to perform my job** | ⬭ | ⬭ | ⬭ | ⬭ | ⬭ | ⬭ |

35. I experience the following emotional repercussions after any such complication: *

*Check all that apply.*

- [ ] Sadness
- [ ] Anxiety
- [ ] Anger
- [ ] Guilt
- [ ] Shame/embarassment
- [ ] Fear of legal action
- [ ] Depression
- [ ] Burnout
- [ ] No "negative" emotions experience
- [ ] Other: ______________________

36. Please specify any additional emotional considerations you have related to reporting intraoperative adverse events:

If any of the above are Not Applicable, please specify below

______________________
______________________
______________________
______________________
______________________

37. Perceived benefits *

*Mark only one oval per row.*

| | 5 = Strongly agree / Very much describes my practice | 4 | 3 = Neither agree nor disagree / this may or may not describe my practice, or my practice has too much variation | 2 | 1 = Strongly disagree / does not describe my practice | N/A |
|---|---|---|---|---|---|---|
| **There is clinical utility** | ◯ | ◯ | ◯ | ◯ | ◯ | ◯ |

| | | | | | | |
|---|---|---|---|---|---|---|
| **to reporting these complications** | ◯ | ◯ | ◯ | ◯ | ◯ | ◯ |
| **There is quality assessment or improvement utility to reporting these complications** | ◯ | ◯ | ◯ | ◯ | ◯ | ◯ |
| **Reporting these complications may enhance the institutional safety culture** | ◯ | ◯ | ◯ | ◯ | ◯ | ◯ |
| **Reporting these complications may enhance patient safety and outcomes** | ◯ | ◯ | ◯ | ◯ | ◯ | ◯ |
| **Reporting these events may add educational value** | ◯ | ◯ | ◯ | ◯ | ◯ | ◯ |

38. Please specify any additional benefits to reporting intraoperative adverse events:

If any of the above are Not Applicable, please specify below

_______________________________________________

_______________________________________________

_______________________________________________

_______________________________________________

_______________________________________________

39. Perceived consequences to clinical practice *

*Mark only one oval per row.*

| | 5 = Strongly agree / Very much describes my practice | 4 | 3 = Neither agree nor disagree / this may or may not describe my practice, or my practice has too much variation | 2 | 1 = Strongly disagree / does not describe my practice | N/A |
|---|---|---|---|---|---|---|
| **Reporting these complications may decrease my likelihood to take any form of risk** | ◯ | ◯ | ◯ | ◯ | ◯ | ◯ |
| **Reporting these complications may decrease the quality of my surgical practice** | ◯ | ◯ | ◯ | ◯ | ◯ | ◯ |

40. Please specify any additional potential consequences that may factor into your decision to report intraoperative adverse events:

If any of the above are Not Applicable, please specify below

PART 5 - Next Steps for Intraoperative Adverse Event (iAE) Reporting

41. Please indicate which of the following you believe would improve or increase intraoperative reporting.. all options that apply)

*Check all that apply.*

| | iAE related professional society guidelines | iAE assessment/grading classification system | iAE assessment/grading forms and tools | Additional financial or admininistrative support | Shift in practice culture surrounding iAE reporting |
|---|---|---|---|---|---|
| **... in your practice** | ☐ | ☐ | ☐ | ☐ | ☐ |
| **... globally** | ☐ | ☐ | ☐ | ☐ | ☐ |

42. Please include any additional comments, suggestions, questions, and concerns related to iAE reportir

#### PART 6 - ICARUS Reporting Guidelines Global Assessment

The aim of the Intraoperative Complication Assessment and Reporting with Universal Standards (ICARUS) criteri is to standardize the collection of intraoperative Adverse Events.

Using a Delphi methodology, supported by an extensive, 4 year-long, umbrella review of studies reporting intraoperative outcomes during surgical procedures, we have developed a standard set of 13 CRITERIA to standardize the assessment, reporting, and grading of iAEs in ALL surgical specialties.

The ICARUS guidelines have the mandate of guiding surgeons, anesthesiologists, and nurses in reporting iAEs accurately.

Herein, we ask you to rate each criterion (scale 1-5) to assist with the GLOBAL ASSESSMENT VALIDATION .

Optionally, you may provide comments after rating each criterion.

43. CRITERION n1. IN A STUDY REPORTING PERIOPERATIVE OUTCOMES, THE INTRAOPERATIVE ADVERSE EVENTS (IAEs) SHOULD BE REPORTED AS ONE OF THE OUTCOMES OF INTEREST

The umbrella review of the studies assessing the iAE complication reporting as one of the outcomes of inter[illegible] showed that approximately only 50% of papers reporting perioperative outcomes assessed iAEs as an outcome of interest. The panel feels that NOT reporting the iAEs as an outcome of interest is not equivalent [illegible] not having any iAEs. In cases where no iAEs occur, the surgeons and authors should state as follows: "no iA[illegible] has occurred"

*Mark only one oval per row.*

| | 5: very useful | 4 | 3 | 2 | 1: not useful |
|---|---|---|---|---|---|
| **CLARITY** | ◯ | ◯ | ◯ | ◯ | ◯ |
| **EXHAUSTIVENESS** | ◯ | ◯ | ◯ | ◯ | ◯ |
| **CLINICAL USEFULNESS** | ◯ | ◯ | ◯ | ◯ | ◯ |
| **QUALITY ASSESSMENT AND IMPROVEMENT PERSPECTIVE** | ◯ | ◯ | ◯ | ◯ | ◯ |
| **RESEARCH UTILITY** | ◯ | ◯ | ◯ | ◯ | ◯ |

44. CRITERION n1. COMMENTS (Optional)

45. CRITERION n2.THE INTRAOPERATIVE ADVERSE EVENTS (IAEs) AND THE DEFINITION OF EACH SPECIFIC INTRAOPERATIVE ADVERSE EVENT SHOULD BE REPORTED OR REFERENCED

The definition of each iAE collected should be provided or referenced in the methods in order to reduce the heterogeneity between studies

*Mark only one oval per row.*

| | 5: very useful | 4 | 3 | 2 | 1: not useful |
|---|---|---|---|---|---|
| **CLARITY** | ◯ | ◯ | ◯ | ◯ | ◯ |
| **EXHAUSTIVENESS** | ◯ | ◯ | ◯ | ◯ | ◯ |
| **CLINICAL USEFULNESS** | ◯ | ◯ | ◯ | ◯ | ◯ |
| **QUALITY ASSESSMENT AND IMPROVEMENT PERSPECTIVE** | ◯ | ◯ | ◯ | ◯ | ◯ |
| **RESEARCH UTILITY** | ◯ | ◯ | ◯ | ◯ | ◯ |

46. CRITERION n2. COMMENTS (Optional)

47. CRITERION n3. EACH INTRAOPERATIVE ADVERSE EVENT (IAE) SHOULD BE REPORTED USING ONE OF THE PROPOSED CLASSIFICATION SYSTEMS (ClassIntra, EAUiaiC, iAE severity classification scheme or modified Satava)

The intraoperative complication classifications are rarely reported. The panel would encourage the use of on of the aforementioned  standardized classifications of the intraoperative complications (ClassIntra, EAUiaiC, iAE severity classification scheme or modified Satava)

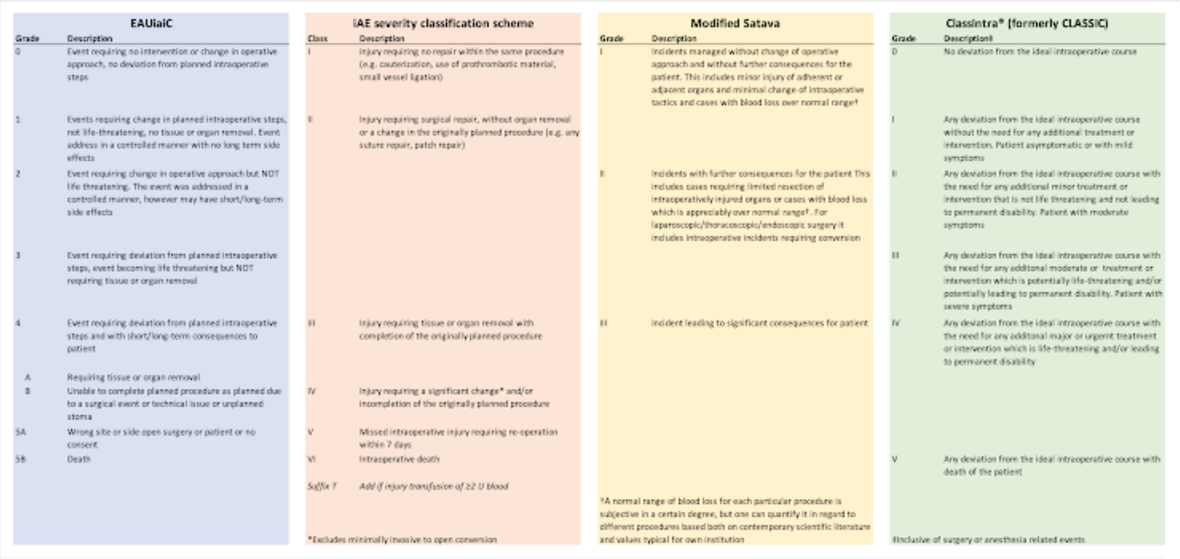

**EAUiaiC**

| Grade | Description |
|---|---|
| 0 | Event requiring no intervention or change in operative approach, no deviation from planned intraoperative steps |
| 1 | Events requiring change in planned intraoperative steps, not life-threatening, no tissue or organ removal. Event address in a controlled manner with no long term side effects |
| 2 | Event requiring change in operative approach but NOT life threatening. The event was addressed in a controlled manner, however may have short/long-term side effects |
| 3 | Event requiring deviation from planned intraoperative steps, event becoming life threatening but NOT requiring tissue or organ removal |
| 4 | Event requiring deviation from planned intraoperative steps and with short/long-term consequences to patient |
| A | Requiring tissue or organ removal |
| B | Unable to complete planned procedure as planned due to a surgical event or technical issue or unplanned stoma |
| 5A | Wrong site or side open surgery or patient or no consent |
| 5B | Death |

**iAE severity classification scheme**

| Class | Description |
|---|---|
| I | Injury requiring no repair within the same procedure (e.g. cauterization, use of prothrombotic material, small vessel ligation) |
| II | Injury requiring surgical repair, without organ removal or a change in the originally planned procedure (e.g. any suture repair, patch repair) |
| III | Injury requiring tissue or organ removal with completion of the originally planned procedure |
| IV | Injury requiring a significant change* and/or incompletion of the originally planned procedure |
| V | Missed intraoperative injury requiring re-operation within 7 days |
| VI | Intraoperative death |
| *Suffix T* | *Add if injury transfusion of ≥2 U blood* |

*Excludes minimally invasive to open conversion

**Modified Satava**

| Grade | Description |
|---|---|
| I | Incidents managed without change of operative approach and without further consequences for the patient. This includes minor injury of adherent or adjacent organs and minimal change of intraoperative tactics and cases with blood loss over normal range† |
| II | Incidents with further consequences for the patient This includes cases requiring limited resection of intraoperatively injured organs or cases with blood loss which is appreciably over normal range†. For laparoscopic/thoracoscopic/endoscopic surgery it includes intraoperative incidents requiring conversion |
| III | Incident leading to significant consequences for patient |

†A normal range of blood loss for each particular procedure is subjective in a certain degree, but one can quantify it in regard to different procedures based both on contemporary scientific literature and values typical for own institution

**ClassIntra* (formerly CLASSIC)**

| Grade | Description‡ |
|---|---|
| 0 | No deviation from the ideal intraoperative course |
| I | Any deviation from the ideal intraoperative course without the need for any additional treatment or intervention. Patient asymptomatic or with mild symptoms |
| II | Any deviation from the ideal intraoperative course with the need for any additional minor treatment or intervention that is not life threatening and not leading to permanent disability. Patient with moderate symptoms |
| III | Any deviation from the ideal intraoperative course with the need for any additonal moderate or treatment or intervention which is potentially life-threatening and/or potentially leading to permanent disability. Patient with severe symptoms |
| IV | Any deviation from the ideal intraoperative course with the need for any additonal major or urgent treatment or intervention which is life-threatening and/or leading to permanent disability |
| V | Any deviation from the ideal intraoperative course with death of the patient |

‡Inclusive of surgery or anesthesia related events

[Click here for enlarged version of classification schemata: https://tinyurl.com/iAE-Grading ]

*Mark only one oval per row.*

| | 5: very useful | 4 | 3 | 2 | 1: not useful |
|---|---|---|---|---|---|
| **CLARITY** | ◯ | ◯ | ◯ | ◯ | ◯ |
| **EXHAUSTIVENESS** | ◯ | ◯ | ◯ | ◯ | ◯ |
| **CLINICAL USEFULNESS** | ◯ | ◯ | ◯ | ◯ | ◯ |
| **QUALITY ASSESSMENT AND IMPROVEMENT PERSPECTIVE** | ◯ | ◯ | ◯ | ◯ | ◯ |
| **RESEARCH UTILITY** | ◯ | ◯ | ◯ | ◯ | ◯ |

48. CRITERION n3. COMMENTS (Optional)

49. CRITERION n4. EACH INTRAOPERATIVE ADVERSE EVENT (IAE) SHOULD BE REPORTED SEPARATELY BY GRADE

When adverse events are reported, it is important to grade them according to the preferrred iAEs classificatic systems outlined in the previous question

*Mark only one oval per row.*

| | 5: very useful | 4 | 3 | 2 | 1: not useful |
|---|---|---|---|---|---|
| **CLARITY** | | | | | |
| **EXHAUSTIVENESS** | | | | | |
| **CLINICAL USEFULNESS** | | | | | |
| **QUALITY ASSESSMENT AND IMPROVEMENT PERSPECTIVE** | | | | | |
| **RESEARCH UTILITY** | | | | | |

50. CRITERION n4. COMMENTS (Optional)

51. CRITERION n5. ANESTHESIOLOGICAL AND SURGICAL COMPLICATIONS SHOULD BE REPORTED SEPARATELY

Anesthesiological and surgical complications have different causes and treatment. A report of both anesthesiological and surrgical iAEs would be recommended

*Mark only one oval per row.*

| | 5: very useful | 4 | 3 | 2 | 1: not useful |
|---|---|---|---|---|---|
| **CLARITY** | ⬭ | ⬭ | ⬭ | ⬭ | ⬭ |
| **EXHAUSTIVENESS** | ⬭ | ⬭ | ⬭ | ⬭ | ⬭ |
| **CLINICAL USEFULNESS** | ⬭ | ⬭ | ⬭ | ⬭ | ⬭ |
| **QUALITY ASSESSMENT AND IMPROVEMENT PERSPECTIVE** | ⬭ | ⬭ | ⬭ | ⬭ | ⬭ |
| **RESEARCH UTILITY** | ⬭ | ⬭ | ⬭ | ⬭ | ⬭ |

52. CRITERION n5. COMMENTS (Optional)

53. CRITERION n6. THE NUMBER OF INTRAOPERATIVE ADVERSE EVENTS AND THE NUMBER OF PATIENTS REPORTING THE INTRAOPERATIVE ADVERSE EVENTS (IAEs) SHOULD BE REPORTED SEPARATELY

The number of patients that report an adverse event may differ from the total number of events. For exampl in a cohort of 100 patients 20 patients (20%) reported a total of 34 events.

*Mark only one oval per row.*

| | 5: very useful | 4 | 3 | 2 | 1: not useful |
|---|---|---|---|---|---|
| **CLARITY** | ◯ | ◯ | ◯ | ◯ | ◯ |
| **EXHAUSTIVENESS** | ◯ | ◯ | ◯ | ◯ | ◯ |
| **CLINICAL USEFULNESS** | ◯ | ◯ | ◯ | ◯ | ◯ |
| **QUALITY ASSESSMENT AND IMPROVEMENT PERSPECTIVE** | ◯ | ◯ | ◯ | ◯ | ◯ |
| **RESEARCH UTILITY** | ◯ | ◯ | ◯ | ◯ | ◯ |

54. CRITERION n6. COMMENTS (Optional)

55. CRITERION n7. WHEN APPROPRIATE, CONDITIONS ASSOCIATED WITH INTRAOPERATIVE ADVERSE EVENTS (IAEs) SHOULD BE REPORTED

Not all iAEs are related to the surgery. Example include some preexisting conditions (e.g. past medical histo of pelvic radiation would lead to a difficult posterior prostate detachment leading to a rectal perforation), atypical anatomy (e.g. atypical vessels variants causing intraoperative bleeding that required prolonged cauterization or vessel suturing), or malfunctioning surgical instruments (e.g. malfunction of the electrocaut or malposition of the protective sheath of the robotic scissors leading to a vessel injury)

*Mark only one oval per row.*

| | 5: very useful | 4 | 3 | 2 | 1: not useful |
|---|---|---|---|---|---|
| **CLARITY** | ◯ | ◯ | ◯ | ◯ | ◯ |
| **EXHAUSTIVENESS** | ◯ | ◯ | ◯ | ◯ | ◯ |
| **CLINICAL USEFULNESS** | ◯ | ◯ | ◯ | ◯ | ◯ |
| **QUALITY ASSESSMENT AND IMPROVEMENT PERSPECTIVE** | ◯ | ◯ | ◯ | ◯ | ◯ |
| **RESEARCH UTILITY** | ◯ | ◯ | ◯ | ◯ | ◯ |

56. CRITERION n7 COMMENTS (Optional)

57. CRITERION n8. IF THE INTRAOPERATIVE ADVERSE EVENT (IAE) REQUIRES A CONVERSION, BOTH THE IAE THAT CAUSED THE CONVERSION AND THE ACTION UNDERTAKEN SHOULD BE REPORTED

Conversion due to iAEs could impact dramatically on the postoperative course and management with appropriate reporting. (e.g. switching from an approach to an alternate approach, from a technique to an alternate technique, or abortion of the procedure)

*Mark only one oval per row.*

| | 5: very useful | 4 | 3 | 2 | 1: not useful |
|---|---|---|---|---|---|
| **CLARITY** | ⬭ | ⬭ | ⬭ | ⬭ | ⬭ |
| **EXHAUSTIVENESS** | ⬭ | ⬭ | ⬭ | ⬭ | ⬭ |
| **CLINICAL USEFULNESS** | ⬭ | ⬭ | ⬭ | ⬭ | ⬭ |
| **QUALITY ASSESSMENT AND IMPROVEMENT PERSPECTIVE** | ⬭ | ⬭ | ⬭ | ⬭ | ⬭ |
| **RESEARCH UTILITY** | ⬭ | ⬭ | ⬭ | ⬭ | ⬭ |

58. CRITERION n8. COMMENTS (Optional)

59. CRITERION n9. THE INTRAOPERATIVE ADVERSE EVENTS (IAEs) SHOULD BE REPORTED SPECIFYING THE SURGICAL STEP THAT WAS ASSOCIATED WITH OR AFFECTED BY THE IAEs

Reporting the surgical steps during which iAEs occur is important and can help surgeons and trainees raise awareness of specific surgical step-dependent complications

*Mark only one oval per row.*

| | 5: very useful | 4 | 3 | 2 | 1: not useful |
|---|---|---|---|---|---|
| **CLARITY** | | | | | |
| **EXHAUSTIVENESS** | | | | | |
| **CLINICAL USEFULNESS** | | | | | |
| **QUALITY ASSESSMENT AND IMPROVEMENT PERSPECTIVE** | | | | | |
| **RESEARCH UTILITY** | | | | | |

60. CRITERION n9. COMMENTS (Optional)

61. CRITERION n10. THE TIMING OF THE THE INTRAOPERATIVE ADVERSE EVENTS (IAEs) ASSESSMENT SHOULD BE REPORTED AS FOLLOWS (see below):

Reporting the surgical steps during which a iAEs occur is important and can help surgeons and trainees raise awareness of specific surgical step-dependent complications

(1) if an iAE is recognized ***during the surgical procedure***, the panel recommend debriefing after the surgical procedure for collecting iAEs that are recognized intraoperatively (using the further on described iAEs PATIENT FORM)

(2) if an iAE is ***not recognized during the surgical procedure,*** report the point at which the iAE became apparent in the postoperative course.

*Mark only one oval per row.*

| | 5: very useful | 4 | 3 | 2 | 1: not useful |
|---|---|---|---|---|---|
| **CLARITY** | | | | | |
| **EXHAUSTIVENESS** | | | | | |
| **CLINICAL USEFULNESS** | | | | | |
| **QUALITY ASSESSMENT AND IMPROVEMENT PERSPECTIVE** | | | | | |
| **RESEARCH UTILITY** | | | | | |

62. CRITERION n10. COMMENTS (Optional)

63. CRITERION n11. THE MANAGEMENT OF THE THE INTRAOPERATIVE ADVERSE EVENTS (IAEs) SHOULD BE REPORTED

Information on the management if an iAES is important and could provide important insight for colleagues th may find themselves dealing with the same iAEs.

*Mark only one oval per row.*

| | 5: very useful | 4 | 3 | 2 | 1: not useful |
|---|---|---|---|---|---|
| **CLARITY** | ◯ | ◯ | ◯ | ◯ | ◯ |
| **EXHAUSTIVENESS** | ◯ | ◯ | ◯ | ◯ | ◯ |
| **CLINICAL USEFULNESS** | ◯ | ◯ | ◯ | ◯ | ◯ |
| **QUALITY ASSESSMENT AND IMPROVEMENT PERSPECTIVE** | ◯ | ◯ | ◯ | ◯ | ◯ |
| **RESEARCH UTILITY** | ◯ | ◯ | ◯ | ◯ | ◯ |

64. CRITERIA n11. COMMENTS (Optional)

65. CRITERION n12. REPORT THE SEQUELAE OF A GIVEN INTRAOPERATIVE ADVERSE EVENTS (IAES) IN THE POSTOPERATIVE COURSE AS FOLLOWS (see below).
NB: THIS NEW ADJUNCTIVE CLASSIFICATION IS COMPATIBLE WITH ALL THE PREVIOUSLY REPORTED CLASSIFICATION FOR IAEs AND IS NOT IN CONFLICT WITH THE CLAVIEN DINDO CLASSIFICATION SYSTEM. IT WILL ADD CLARITY IN THE ASSESSMENT OF EACH iAE AND ITS IMPACT ON THE POSTOPERATIVE COURSE

An iAE may impact the postoperative course. It could be related to postoperative complications and to postoperative sequelae that will be permanent, impacting patient quality of life (e.g. obturator neuropathies a serious complications associated with RALP. These complications are usually a direct result of physical nerv damage intraoperatively, which will present clinically with motor and sensory deficits to the lower limb; althou uncommon, most neuropathies that develop as a result of intraoperative injury resolve within a few weeks unless complete transection or ligation occurs)

(a) without postoperative sequelae

(b) with non-permanent postoperative sequelae

(c) with permanent postoperative sequelae

(d) requiring a re-operation (immediate or late post-op, or after discharge)

(e) postoperative death

*Mark only one oval per row.*

| | 5: very useful | 4 | 3 | 2 | 1: not useful |
|---|---|---|---|---|---|
| **CLARITY** | | | | | |
| **EXHAUSTIVENESS** | | | | | |
| **CLINICAL USEFULNESS** | | | | | |
| **QUALITY ASSESSMENT AND IMPROVEMENT PERSPECTIVE** | | | | | |
| **RESEARCH UTILITY** | | | | | |

66. CRITERION n12. COMMENTS (Optional)

67. CRITERION n13. REPORT CHANGES TO THE CLINICAL COURSE THAT WERE ASSOCIATED WITH ANY INTRAOPERATIVE ADVERSE EVENTS (IAEs)

In addition to reporting patient-centric sequelae, it is important to report changes to the clinical course that resulted from the iAEs, such as operative time (either increase for management or decrease due to abortion operation), extension in hospitalization duration, additional procedures, ICU stay, or unplanned specialty consults.

*Mark only one oval per row.*

| | 5: very useful | 4 | 3 | 2 | 1: not useful |
|---|---|---|---|---|---|
| **CLARITY** | | | | | |
| **EXHAUSTIVENESS** | | | | | |
| **CLINICAL USEFULNESS** | | | | | |
| **QUALITY ASSESSMENT AND IMPROVEMENT PERSPECTIVE** | | | | | |
| **RESEARCH UTILITY** | | | | | |

68. CRITERION n13. COMMENTS (Optional)

69. ANY OTHER CRITERIA THAT YOU FEEL ARE IMPORTANT TO INCLUDE (OPTIONAL OPEN QUESTION):

#### Part 7 - Intraoperative Adverse Events (iAEs) Reporting Resources and Tools

##### Data Collection

70. In terms of data collection, how useful or helpful would the following be for properly assessing, grading and reporting iAEs

*Mark only one oval per row.*

| | 5: very useful | 4 | 3 | 2 | 1: not useful |
|---|---|---|---|---|---|
| **Patient form** | ⬭ | ⬭ | ⬭ | ⬭ | ⬭ |
| **Online grade calculator/converter** | ⬭ | ⬭ | ⬭ | ⬭ | ⬭ |
| **Automated data recording** | ⬭ | ⬭ | ⬭ | ⬭ | ⬭ |
| **Post-operative time-out or checklist** | ⬭ | ⬭ | ⬭ | ⬭ | ⬭ |

71. Additional resources that may be helpful for data collection (Optional)

#### Scientific Publication

72. In terms of scientific publication, how useful or helpful would the following be for properly assessing, grading, and reporting iAEs

*Mark only one oval per row.*

| | 5: very useful | 4 | 3 | 2 | 1: not useful |
|---|---|---|---|---|---|
| **Criteria checklist** | ⬭ | ⬭ | ⬭ | ⬭ | ⬭ |

73. In order to standardize iAE reporting in scientific literature, how important is it that academic journals offer guideline recommendations for properly assessing, grading, and reporting iAEs?

*Mark only one oval per row.*

| | 5: very important | 4 | 3 | 2 | 1: not important |
|---|---|---|---|---|---|
| **Importance** | ⬭ | ⬭ | ⬭ | ⬭ | ⬭ |

74. Additional resources that may be helpful for scientific publication (Optional)

_______________________________________________

_______________________________________________

_______________________________________________

_______________________________________________

_______________________________________________

#### Thank you

Thank you for taking the time to complete this survey. If you are done, you may submit your responses.

#### Acknowledgement (Optional)

If you would like your name to be included in a future publication of the ICARUS, please complete the below questic prior to survey submission.

75. First Name and Middle Initial

______________________________

76. Last Name

______________________________

77. Institution

______________________________

78. City

______________________________

79. Email

______________________________

This content is neither created nor endorsed by Google.

Google Forms

Dear Colleague,

Thank you for taking part in the 1st and 2nd Delphi rounds on the **"Intraoperative Adverse Events Definition" Delphi Consensus** (over 4000 participants worldwide).

This is the 3**rd Delphi round** of the ICARUS Global Survey, and the questions will take you no longer than **2 minutes to complete.**

You may open the survey in your web browser by clicking the link below:
ICARUS Adverse Event Definitions (New link!)

This link should not be forwarded to others.

Please fill out the forms at your earliest convenience by **September 20th, 2022**

***Only those who complete the short survey and confirm the AUTHORSHIP details at the end of the survey (Name, Last Name, and Institution) will be listed as part of the collaborative authorship in the future publication of the results of this Delphi Survey.***

Thank you,

Dr. Giovanni Cacciamani MD, MSc, FEBU
University of Southern California (USC)

On behalf of ICARUS Global Surgical Collaboration Project

### ICARUS Adverse Event Definitions

Thank you for your gracious participation in the first ICARUS survey.

As part of a modified Delphi consensus survey, we would like to invite you to complete ROUND 3 of the Delphi Survey the following 2-minute survey as a follow-up to the initial ICARUS survey.

You will be presented with 7 definitions for adverse events and you will be asked to rate your agreement with each definition on a scale from 1-5.

All those who complete the short survey in its entirety will be listed as part of the collaborative authorship in the future publication of the results of this Delphi Survey.

#### Adverse Event Definitions

**In the following sections, you will be presented with adverse event definitions by timing relative to procedure and by level of anesthesia. Please indicate your level of agreement with the definitions.**

#### Definition of PREOPERATIVE ADVERSE EVENT

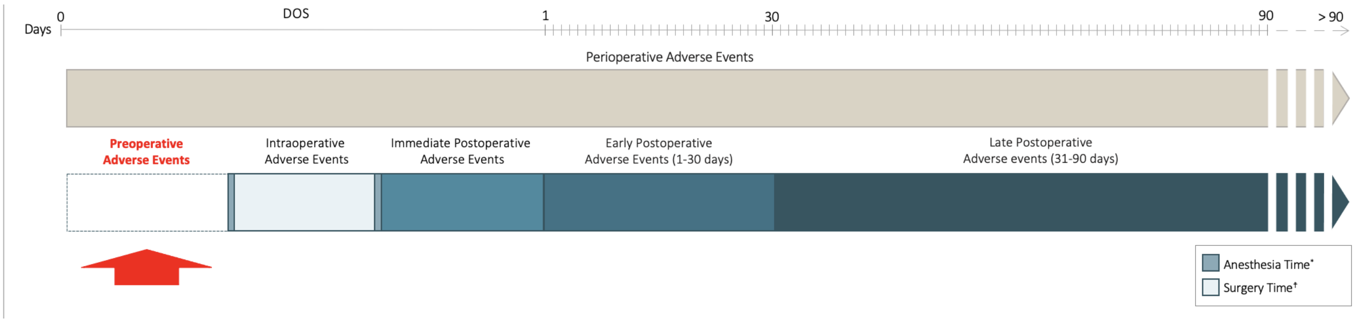


A PREOPERATIVE ADVERSE EVENT is any unintended, unplanned, and possibly anticipable/preventable surgical/interventional and/or anesthesiologic and/or nursing incident, potentially resulting in a complication, which occurs prior to anesthesia1 or surgical/interventional procedure time2, but while the patient is in the pre-op room or procedure/surgery room (e.g., patient positioning, disinfection, prophylactic medication administration, etc.), and which might be harmful to the patient, regardless of whether the event is recognized during or after the preoperative period.

Note 1: Anesthesia Time is a continuous time period from the start of anesthesia to the end of an anesthesia service (as defined by 2019 ASA RVG).

Note 2: Surgical/Interventional Procedure Time is defined as the time period from when the initial incision was made for the principal procedure to skin closure. In the case of endoscopic maneuvers which do not require any skin incision (i.e.: cystoscopy, colonoscopy, etc.) the surgical procedure time is defined as the time period from when the endoscopic tool is initially inserted for the principal procedure through the natural or pre-existing orifice to its withdrawal.)

Delphi Round 2 definition agreement: 93.7%

Agreement with definition

◯ 5 Strongly agree
◯ 4
◯ 3 Neither agree nor disagree
◯ 2
◯ 1 Strongly disagree

Comment regarding definition of preoperative adverse events (Optional)

__________________________________

Comment regarding definition of preoperative adverse events (Required)

__________________________________

#### Definition of INTRAOPERATIVE ADVERSE EVENT (IAE)

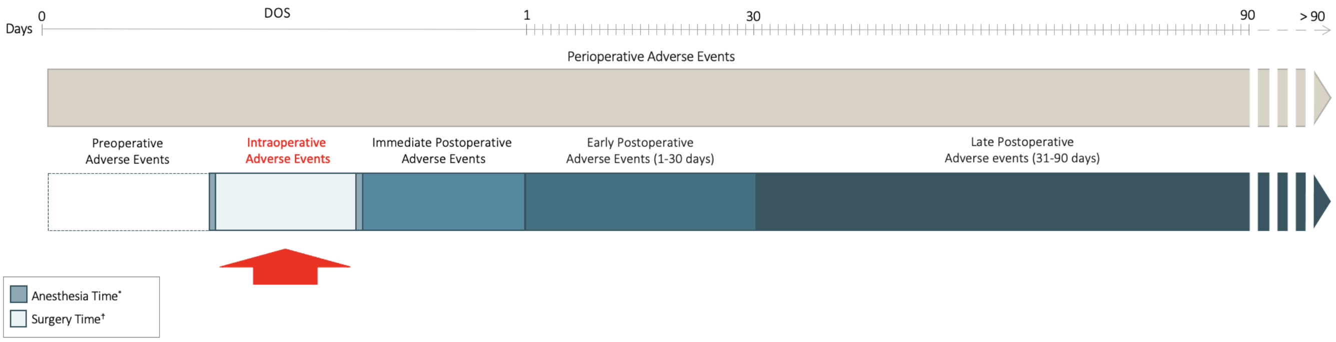


An INTRAOPERATIVE ADVERSE EVENT is any unintended, unplanned, and possibly anticipable/preventable surgical/interventional and/or anesthesiologic and/or nursing incident leading to any deviation from the ideal procedure plana, potentially resulting in a complication, which occurs during anesthesia1 and/or surgical/interventional procedure time2, and which may be harmful to the patient, regardless of whether the event is recognized during or after the intraoperative period.

Note a: "Ideal"  is used to be consistent with previous publications: Cunningham et al. World J Surg (2009) 33:1099-1100 and Clavien et al. World J Surg (2008) 32:939-941

Note 1: Anesthesia Time is a continuous time period from the start of anesthesia to the end of an anesthesia service (as defined by 2019 ASA RVG).

Note 2: Surgical/Interventional Procedure Time is defined as the time period from when the initial incision was made for the principal procedure to skin closure. In the case of endoscopic maneuvers which do not require any skin incision (i.e.: cystoscopy, colonoscopy, etc.) the surgical procedure time is defined as the time period from when the endoscopic tool is initially inserted for the principal procedure through the natural or pre-existing orifice to its withdrawal.)

Delphi Round 2 definition agreement: 96.9%

Agreement with definition

◯ 5 Strongly agree
◯ 4
◯ 3 Neither agree nor disagree
◯ 2
◯ 1 Strongly disagree

Comments regarding the definition for iAEs (Optional)

__________________________________

Comments regarding the definition for iAEs (Required)

__________________________________

#### Definition of iAEs under GENERAL ANESTHESIA

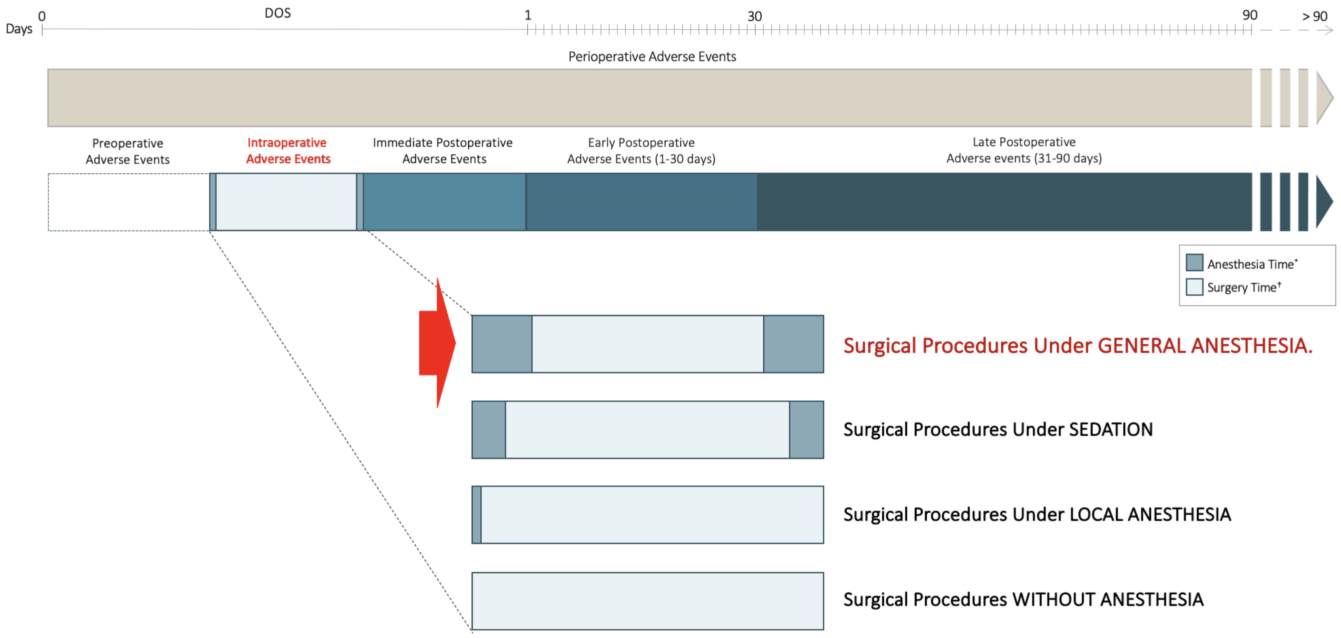


In cases under GENERAL ANESTHESIA, an intraoperative adverse event is any unintended, unplanned, and possibly anticipable/preventable surgical/interventional and/or anesthesiologic and/or nursing incident leading to any deviation from the ideal procedure plana, potentially resulting in a complication, which occurs during anesthesia time1 -- including the entire surgical/interventional procedure time2 -- and which may be harmful to the patient, regardless of whether the event is recognized during or after the intraoperative period.

Note a: "Ideal"  is used to be consistent with previous publications: Cunningham et al. World J Surg (2009) 33:1099-1100 and Clavien et al. World J Surg (2008) 32:939-941

Note 1: Anesthesia Time is a continuous time period from the start of anesthesia to the end of an anesthesia service (as defined by 2019 ASA RVG).

Note 2: Surgical/Interventional Procedure Time is defined as the time period from when the initial incision was made for the principal procedure to skin closure. In the case of endoscopic maneuvers which do not require any skin incision (i.e.: cystoscopy, colonoscopy, etc.) the surgical procedure time is defined as the time period from when the endoscopic tool is initially inserted for the principal procedure through the natural or pre-existing orifice to its withdrawal.)

Delphi Round 2 definition agreement: 96.8%

Agreement with definition

◯ 5 Strongly agree
◯ 4
◯ 3 Neither agree nor disagree
◯ 2
◯ 1 Strongly disagree

Comment on iAEs under general anesthesia (Optional)

__________________________________

Definition of iAEs under general anesthesia (Required

#### Definition of iAEs under SEDATION

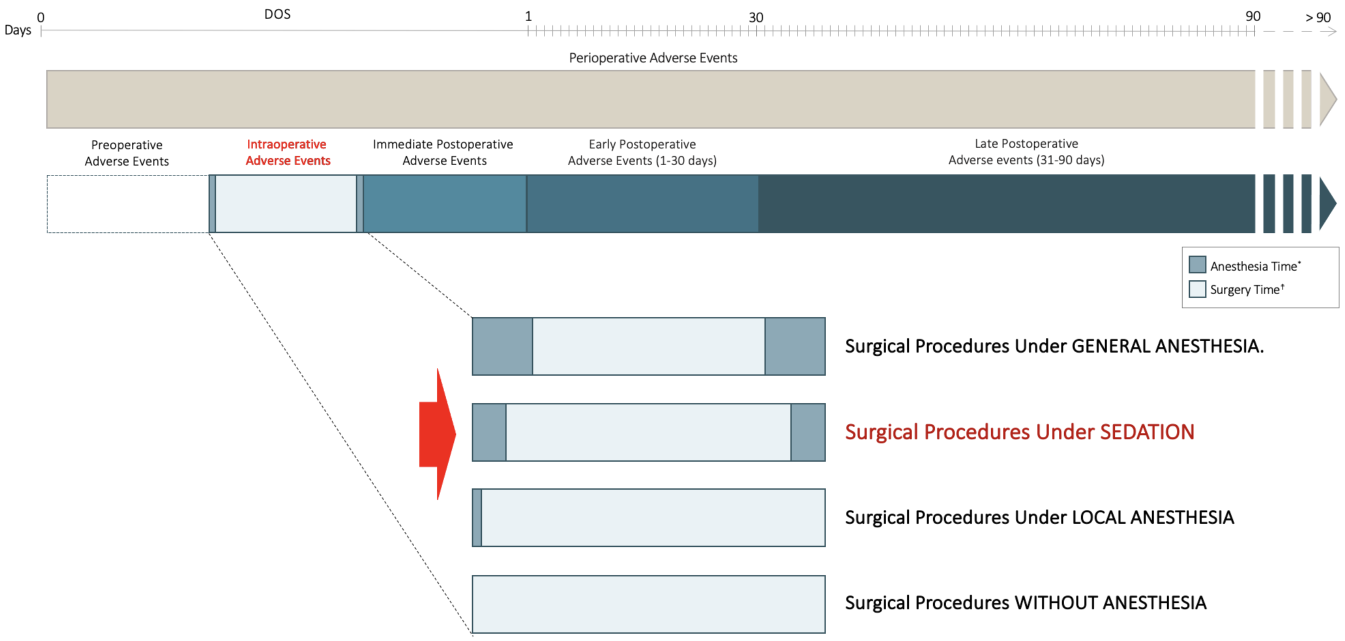


In cases under SEDATION, an intraoperative adverse event is any unintended, unplanned, and possibly anticipable/preventable surgical/interventional and/or anesthesiologic and/or nursing incident leading to any deviation from the ideal procedure plana, potentially resulting in a complication, which occurs during anesthesia time1 -- including the entire surgical/interventional procedure time2 -- and which may be harmful to the patient, regardless of whether the event is recognized during or after the intraoperative period.

Note a: "Ideal"  is used to be consistent with previous publications: Cunningham et al. World J Surg (2009) 33:1099-1100 and Clavien et al. World J Surg (2008) 32:939-941

Note 1: Anesthesia Time is a continuous time period from the start of anesthesia to the end of an anesthesia service (as defined by 2019 ASA RVG).

Note 2: Surgical/Interventional Procedure Time is defined as the time period from when the initial incision was made for the principal procedure to skin closure. In the case of endoscopic maneuvers which do not require any skin incision (i.e.: cystoscopy, colonoscopy, etc.) the surgical procedure time is defined as the time period from when the endoscopic tool is initially inserted for the principal procedure through the natural or pre-existing orifice to its withdrawal.)

Delphi Round 2 definition agreement: 96.6%

Agreement with definition

◯ 5 Strongly agree
◯ 4
◯ 3 Neither agree nor disagree
◯ 2
◯ 1 Strongly disagree

Comments regarding the definition for iAEs under sedation (Optional)

__________________________________

Comments regarding the definition for iAEs under sedation (Required)

#### Definition of iAEs under LOCAL ANESTHESIA

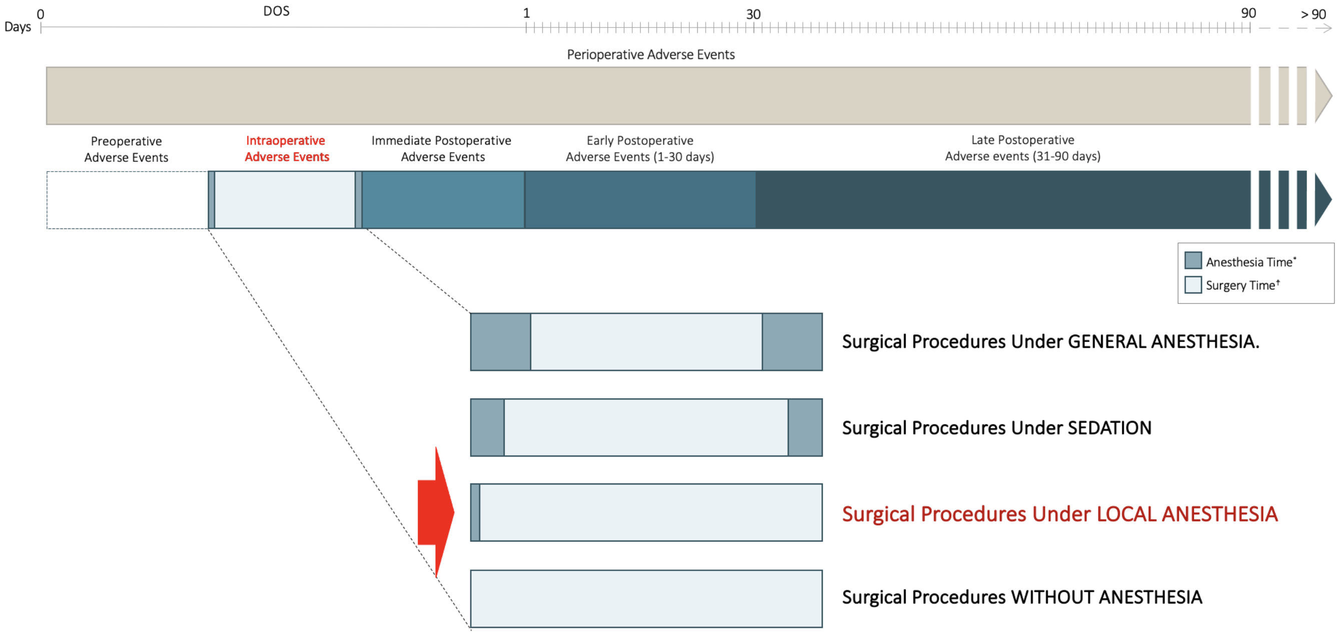


In cases under LOCAL ANESTHESIA, an intraoperative adverse event is any unintended, unplanned, and possibly anticipable/preventable surgical/interventional and/or anesthesiologic and/or nursing incident leading to any deviation from the ideal procedure plana, potentially resulting in a complication, which occurs from the administration of the local anesthetic to the end of the surgical/interventional procedure time2, and which may be harmful to the patient, regardless of whether the event is recognized during or after the intraoperative period.

Note a: "Ideal"  is used to be consistent with previous publications: Cunningham et al. World J Surg (2009) 33:1099-1100 and Clavien et al. World J Surg (2008) 32:939-941

Note 1: Anesthesia Time is a continuous time period from the start of anesthesia to the end of an anesthesia service (as defined by 2019 ASA RVG).

Note 2: Surgical/Interventional Procedure Time is defined as the time period from when the initial incision was made for the principal procedure to skin closure. In the case of endoscopic maneuvers which do not require any skin incision (i.e.: cystoscopy, colonoscopy, etc.) the surgical procedure time is defined as the time period from when the endoscopic tool is initially inserted for the principal procedure through the natural or pre-existing orifice to its withdrawal.)

Delphi Round 2 definition agreement: 96.4%

Agreement with definition

◯ 5 Strongly agree
◯ 4
◯ 3 Neither agree nor disagree
◯ 2
◯ 1 Strongly disagree

Comments regarding the definition for iAEs with local anesthesia (Optional)

______________________________

Comments regarding the definition for iAEs with local anesthesia (Required)

__________________________________________

#### Definition of iAEs WITHOUT ANESTHESIA

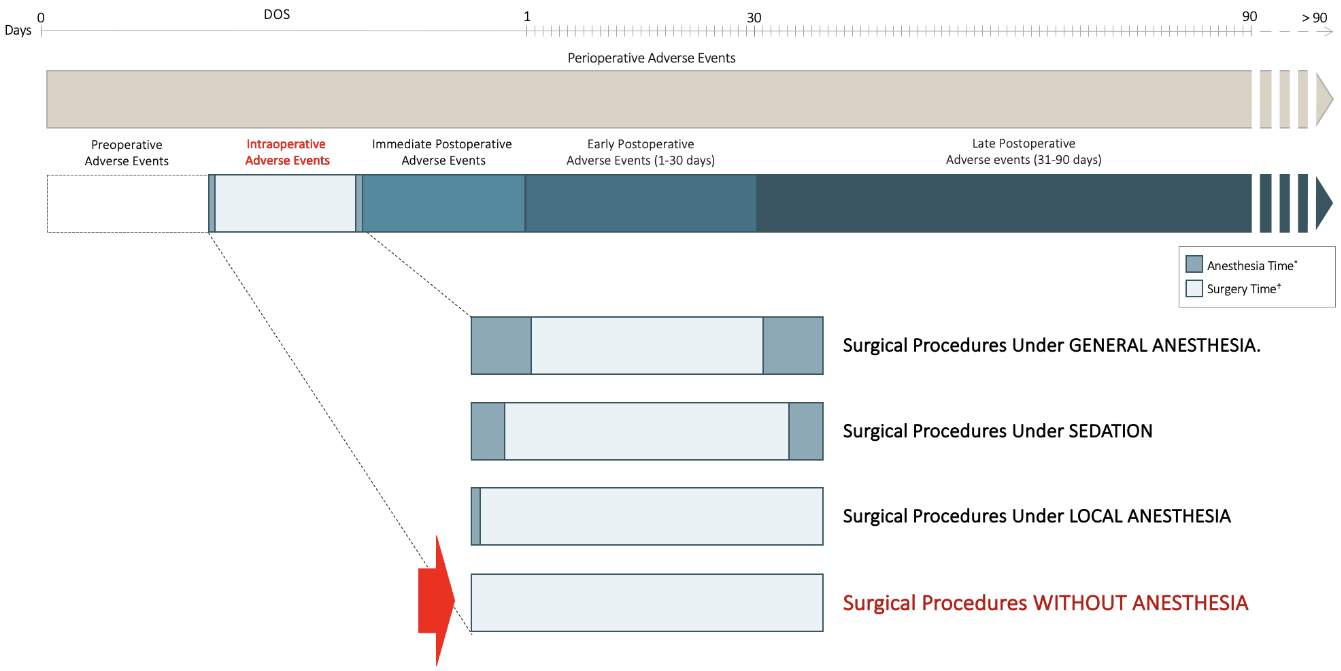


In cases WITHOUT ANESTHESIA, an intraoperative adverse event is any unintended, unplanned, and possibly anticipable/preventable surgical/interventional and/or nursing incident leading to any deviation from the ideal procedure plana, potentially resulting in a complication, which occurs during surgical/interventional procedure time2, and which may be harmful to the patient, regardless of whether the event is recognized during or after the intraoperative period.

Note a: "Ideal"  is used to be consistent with previous publications: Cunningham et al. World J Surg (2009) 33:1099-1100 and Clavien et al. World J Surg (2008) 32:939-941

Note 1: Anesthesia Time is a continuous time period from the start of anesthesia to the end of an anesthesia service (as defined by 2019 ASA RVG).

Note 2: Surgical/Interventional Procedure Time is defined as the time period from when the initial incision was made for the principal procedure to skin closure. In the case of endoscopic maneuvers which do not require any skin incision (i.e.: cystoscopy, colonoscopy, etc.) the surgical procedure time is defined as the time period from when the endoscopic tool is initially inserted for the principal procedure through the natural or pre-existing orifice to its withdrawal.)

Delphi Round 2 definition agreement: 96.5%

Agreement with definition

◯ 5 Strongly agree
◯ 4
◯ 3 Neither agree nor disagree
◯ 2
◯ 1 Strongly disagree

Comments regarding the definition for iAEs without anesthesia (Optional)

______________________________________

Comments regarding the definition for iAEs without anesthesia (Required)

______________________________________

#### Definition of IMMEDIATE POSTOPERATIVE ADVERSE EVENT

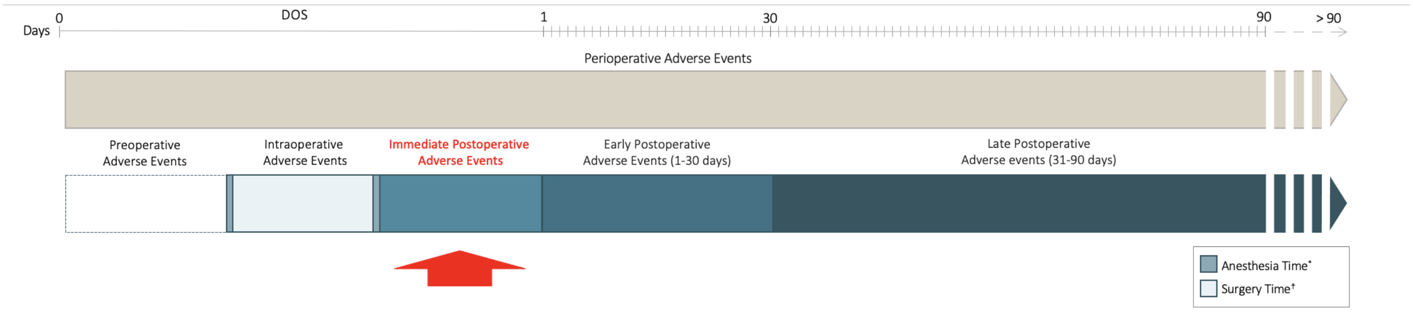


An IMMEDIATE POSTOPERATIVE ADVERSE EVENT is any unintended, unplanned, and possibly anticipable/preventable surgical/interventional and/or anesthesiologic and/or nursing incident leading to any deviation from the ideal procedure plana, potentially resulting in a complication, which occurs after the anesthesia1 and/or surgical/interventional procedure time2 and before postoperative day one (defined as 24 hours after the end of anesthesia time1 -- including the entire surgical/interventional procedure time2 ), and which may be harmful to the patient.

Note a: "Ideal" is used to be consistent with previous publications: Cunningham et al. World J Surg (2009) 33:1099-1100 and Clavien et al. World J Surg (2008) 32:939-941

Note 1: Anesthesia Time is a continuous time period from the start of anesthesia to the end of an anesthesia service (as defined by 2019 ASA RVG).

Note 2: Surgical/Interventional Procedure Time is defined as the time period from when the initial incision was made for the principal procedure to skin closure. In the case of endoscopic maneuvers which do not require any skin incision (i.e.: cystoscopy, colonoscopy, etc.) the surgical procedure time is defined as the time period from when the endoscopic tool is initially inserted for the principal procedure through the natural or pre-existing orifice to its withdrawal.)

Delphi Round 2 definition agreement: 92.5%

Agreement with definition

◯ 5 Strongly agree
◯ 4
◯ 3 Neither agree nor disagree
◯ 2
◯ 1 Strongly disagree

Comment regarding definition of postoperative adverse events (Optional) __________________________

Comment regarding definition of postoperative adverse events (Required) __________________________

#### Collaborative Authorship Information

Last Name

__________________________________

First Name

__________________________________

Degree

__________________________________
((e.g. M.D., M.Sc.))

Department

__________________________________
((Department))

Institution

__________________________________
((Institution))

City

__________________________________
((City))

State

__________________________________
((State))

Country

__________________________________
((Country))

Email

__________________________________
((E-mail address))

Twitter Handle if any (optional)

__________________________________
((Twitter Handle))

Dear Colleague,

As part of the **ICARUS Global Surgical Collaborative Project** community, we need your expertise and invite you to collaborate with us on this new project.
The purpose of this study is to evaluate the inter-rater reliability of a series of intraoperative adverse event (iAE) grading systems (clinicaltrial.gov NCT05270603).

You may open the survey in your web browser by clicking the link below:
INTER-RATER RELIABILITY OF THE INTRAOPERATIVE COMPLICATION CLASSIFICATION SYSTEMS

If the link above does not work, try copying the link below into your web browser:
LiNK

This link is unique to you and should not be forwarded to others.
You will be asked to read and **evaluate intraoperative event scenarios utilizing five intraoperative adverse event grading systems**. There are no additional requirements for participation in this study beyond completing the survey.
In total, the survey should take no more than 10-15 minutes. The deadline to complete the survey is **11.59 pm PST, 20 March 2022.**

After completing the entire survey, you will be given the opportunity to submit your information for **inclusion in the collaborative authorship of our future manuscript.**

Regards,

The ICARUS Global Surgical Collaboration

### INTER-RATER RELIABILITY OF THE INTRAOPERATIVE COMPLICATION CLASSIFICATION SYSTEMS

You are invited to participate in a research study. Your participation is voluntary. This document explains information about this study. You should ask questions about anything that is unclear to you.

Everyone who completes the entire survey will be given the opportunity to submit their information for inclusion in the collaborative authorship.

PURPOSE

The purpose of this study is to evaluate the reliability of a series of intraoperative adverse event (iAE) grading systems. You are invited as a possible participant because of your surgical or procedural experience.

PARTICIPANT INVOLVEMENT

Participants will be asked to read and evaluate intraoperative event scenarios using 5 intraoperative adverse event grading systems. There are no additional requirements for participation in this study beyond completion of the survey. In total, the survey should take no more than 15-20 minutes.

PAYMENT/COMPENSATION FOR PARTICIPATION

Participation in this study is voluntary and there is no compensation for participation in this study.

ALTERNATIVES TO PARTICIPATION

You have the option to not participate in this study.

CONFIDENTIALITY

The members of the research team and the University of Southern California Institutional Review Board (IRB) may access the data. The IRB reviews and monitors research studies to protect the rights and welfare of research subjects.

Data collected in the survey will remain anonymous. No effort will be made to identify or contact participants.

If any personally identifiable information is found in the survey, it will immediately be removed by the study personnel and will not be stored.

INVESTIGATOR CONTACT INFORMATION

If you have any questions about this study, please contact the primary investigator Giovanni Cacciamani, MD by

IRB CONTACT INFORMATION

If you have any questions about your rights as a research participant, please contact the University of Southern California Institutional Review Board at (323) 442-0114 or.

**The following 10 scenarios were selected at random from a list of iAE grading system examples or validation scenarios. (Note: these examples may vary in formatting depending on their original source)**

**For each scenario, you will be asked to respond to the following:**
**A) A series of 15 yes or no questions**
**B) iAE grade using each of the 5 iAE grading systems**

**Suvey responses will remain anonymous.**

**At the end of the survey, you will have the opportunity to submit your information for inclusion as a collaborative author for the associated manuscript.**

Clicking "Next" indicates consent to participation in this study.

#### Surgical experience

Please indicate your level of training

- ◯ Independently practicing physician
- ◯ Resident, fellow, or trainee
- ◯ Other

Please specify training level: ______________________________

Please indicate your specialty:

- ◯ Anesthesiology
- ◯ Cardiothoracic surgery
- ◯ Colon and rectal surgery
- ◯ General surgery
- ◯ Gynecologic oncology
- ◯ Gynecology and obstetrics
- ◯ Neurological surgery
- ◯ Ophthalmologic surgery
- ◯ Oral and maxillofacial surgery
- ◯ Orthopaedic surgery
- ◯ Otorhinolaryngology
- ◯ Pediatric surgery
- ◯ Plastic surgery
- ◯ Surgical oncology
- ◯ Transplant surgery
- ◯ Trauma surgery
- ◯ Urology
- ◯ Vascular surgery
- ◯ Interventional Radiology
- ◯ Interventional Cardiologist
- ◯ Other

Please specify your specialty: ______________________________

| | |
|---|---|
| Which intraoperative adverse event grading system(s) do you typically use? | ☐ I document events without grading<br>☐ iAE Severity<br>☐ Modified Satava<br>☐ EAUiaiC<br>☐ ClassIntra (Formerly CLASSIC)<br>☐ EAES Classification<br>☐ Other |
| Please specify iAE grading system: | ______________________________ |

**SCENARIO 1:**

**Stapling of the superior pulmonary vein during inferior lobectomy, requiring pulmonectomy**

Patient

| | |
|---|---|
| Was the iAE associated with death of the patient?: | ◯ Yes<br>◯ No<br>(If unknown, please select "No") |
| Was the iAE immediately life-threatening?: | ◯ Yes<br>◯ No<br>(If unknown, please select "No") |
| Were there significant consequences to the patient as a result of the iAE?: | ◯ Yes<br>◯ No<br>(If unknown, please select "No". Some examples of iAEs with significant consequences includes injury to or unplanned removal of an otherwise healthy major organ (e.g. unplanned pulmonectomy, nephrectomy) and events that are exceedingly challenging to manage in a controlled manner without potential for long-term patient consequences (e.g. increased risk of postoperative multiorgan failure with major blood transfusion).) |
| Was the incorrect site, side, or surgical approach used without consent?: | ◯ Yes<br>◯ No<br>(If unknown, please select "No") |

Procedure

| | |
|---|---|
| Were there any changes in the ideal intraoperative course related to iAE?: | ◯ Yes<br>◯ No<br>(If unknown, please select "No". Changes in course include minor incidents such as unintended cauterization, equipment malfunction, unanticipated anesthesiologic challenges, or procedural delays regardless of consequence or management.) |
| Was there an unanticipated conversion of approach or significant change to the operative steps of the originally planned procedure due to iAE?: | ◯ Yes<br>◯ No<br>(If unknown, please select "No") |
| Was planned procedure aborted or incomplete due to iAE?: | ◯ Yes<br>◯ No<br>(If unknown, please select "No") |
| Unplanned stoma as a result of iAE?: | ◯ Yes<br>◯ No<br>(If unknown, please select "No") |
| Unplanned tissue or organ removal as a result of iAE?: | ◯ Yes<br>◯ No<br>(If unknown, please select "No") |

iAE Management

| | |
|---|---|
| Was any surgical repair, medical treatment, or other intervention required?: | ◯ Yes<br>◯ No<br>(If unknown, please select "No". Standard procedural management (e.g. cauterization, use of prothombotic material, small vessel ligation) does not qualify as "repair.") |
| Was there a change in post-operative care due to the iAE?: | ◯ Yes<br>◯ No<br>(If unknown, please select "No") |
| Did the iAE or its management necessitate intensive care admission?: | ◯ Yes<br>◯ No<br>(If unknown, please select "No") |
| Was the intraoperative injury missed, necessitating re-operation within 7 days of index procedure?: | ◯ Yes<br>◯ No<br>(If unknown, please select "No") |
| Bleeding Related | |
| Was blood loss appreciably over normal range for procedure?: | ◯ Yes<br>◯ No<br>(If unknown, please select "No". Per Kazaryan, et al. (2013): "A normal range of blood loss for each particular procedure is subjective in a certain degree, but one can quantify it in regard to different procedures based both on contemporary scientific literature and values typical for own institution") |
| Were 2 or more units of blood products required to manage iAE?: | ◯ Yes<br>◯ No<br>(If unknown, please select "No") |
| iAE Grading | |

| | |
|---|---|
| EAUiaiC | ◯ Grade 0: Event requiring no intervention or change in operative approach, no deviation from planned intraoperative steps<br>◯ Grade 1: Events requiring change in planned intraoperative steps, not life-threatening, no tissue or organ removal. Event address in a controlled manner with no long term side effects<br>◯ Grade 2: Event requiring change in operative approach but NOT life threatening. The event was addressed in a controlled manner, however may have short/long-term side effects<br>◯ Grade 3: Event requiring deviation from planned intraoperative steps, event becoming life threatening but NOT requiring tissue or organ removal<br>◯ Grade 4: Event requiring deviation from planned intraoperative steps and with short/long-term consequences to patient<br>◯ Grade 4A: Requiring tissue or organ removal<br>◯ Grade 4B: Unable to complete planned procedure as planned due to a surgical event or technical issue or unplanned stoma<br>◯ Grade 5A: Wrong site or side open surgery or patient or no consent<br>◯ Grade 5B: Death |
| iAE Severity Classification Scheme<br><br>*Excludes minimally invasive to open conversion | ◯ Class I: Injury requiring no repair within the same procedure (e.g. cauterization, use of prothrombotic material, small vessel ligation)<br>◯ Class II: Injury requiring surgical repair, without organ removal or a change in the originally planned procedure (e.g. any suture repair, patch repair)<br>◯ Class III: Injury requiring tissue or organ removal with completion of the originally planned procedure<br>◯ Class IV: Injury requiring a significant change* and/or incompletion of the originally planned procedure<br>◯ Class V: Missed intraoperative injury requiring re-operation within 7 days<br>◯ Class IV: Intraoperative death |
| iAE Severity Classification Scheme<br><br>Suffix T<br><br>(Add if injury transfusion of ≥2 U blood) | ◯ Yes<br>◯ No |

| | |
|---|---|
| Modified Satava<br><br>†A normal range of blood loss for each particular procedure is subjective in a certain degree, but one can quantify it in regard to different procedures based both on contemporary scientific literature and values typical for own institution | ◯ Grade I: Incidents managed without change of operative approach and without further consequences for the patient. This includes minor injury of adherent or adjacent organs and minimal change of intraoperative tactics and cases with blood loss over normal range†<br>◯ Grade II: Incidents with further consequences for the patient This includes cases requiring limited resection of intraoperatively injured organs or cases with blood loss which is appreciably over normal range†. For laparoscopic/thoracoscopic/endoscopic surgery it includes intraoperative incidents requiring conversion<br>◯ Grade III: Incident leading to significant consequences for patient |
| ClassIntra ® (formerly CLASSIC)<br><br>‡Inclusive of surgery or anesthesia related events | ◯ Grade 0: No deviation from the ideal intraoperative course<br>◯ Grade I: Any deviation from the ideal intraoperative course without the need for any additional treatment or intervention. Patient asymptomatic or with mild symptoms<br>◯ Grade II: Any deviation from the ideal intraoperative course with the need for any additional minor treatment or intervention that is not life threatening and not leading to permanent disability. Patient with moderate symptoms<br>◯ Grade III: Any deviation from the ideal intraoperative course with the need for any additional moderate or treatment or intervention which is potentially life-threatening and/or potentially leading to permanent disability. Patient with severe symptoms<br>◯ Grade IV: Any deviation from the ideal intraoperative course with the need for any additional major and urgent treatment or intervention. Patient with life threatening symptoms or leading to permanent disability<br>◯ Grade V: Any deviation from the ideal intraoperative course with death of the patient |
| EAES Classification | ◯ Grade 1: Minor error, no damage or corrective action required<br>◯ Grade 2: Minor consequential error requiring correc- tive action but no change in post-operative care<br>◯ Grade 3: Consequential error requiring major correc- tive action and/or change in post-operative pathway<br>◯ Grade 4: Life-threatening complication that requires major or immediate corrective action which led to a significant alteration of the post-operative pathway which may include re-operation or intensive care admission<br>◯ Grade 5: Major consequential error resulting in death |

**SCENARIO 2:**

**Open right radical nephrectomy with an IVC thrombus. Previous open right hemicolectomy, laparoscopic hysterectomy, peptic ulcer perforation repair.**
**Complication/Management: bowel injury. Resection and bowel anastomosis and completion of the nephrectomy**

Patient

| | |
|---|---|
| Was the iAE associated with death of the patient?: | ◯ Yes<br>◯ No<br>(If unknown, please select "No") |
| Was the iAE immediately life-threatening?: | ◯ Yes<br>◯ No<br>(If unknown, please select "No") |
| Were there significant consequences to the patient as a result of the iAE?: | ◯ Yes<br>◯ No<br>(If unknown, please select "No". Some examples of iAEs with significant consequences includes injury to or unplanned removal of an otherwise healthy major organ (e.g. unplanned pulmonectomy, nephrectomy) and events that are exceedingly challenging to manage in a controlled manner without potential for long-term patient consequences (e.g. increased risk of postoperative multiorgan failure with major blood transfusion).) |
| Was the incorrect site, side, or surgical approach used without consent?: | ◯ Yes<br>◯ No<br>(If unknown, please select "No") |

Procedure

| | |
|---|---|
| Were there any changes in the ideal intraoperative course related to iAE?: | ◯ Yes<br>◯ No<br>(If unknown, please select "No". Changes in course include minor incidents such as unintended cauterization, equipment malfunction, unanticipated anesthesiologic challenges, or procedural delays regardless of consequence or management.) |
| Was there an unanticipated conversion of approach or significant change to the operative steps of the originally planned procedure due to iAE?: | ◯ Yes<br>◯ No<br>(If unknown, please select "No") |
| Was planned procedure aborted or incomplete due to iAE?: | ◯ Yes<br>◯ No<br>(If unknown, please select "No") |
| Unplanned stoma as a result of iAE?: | ◯ Yes<br>◯ No<br>(If unknown, please select "No") |

| | |
|---|---|
| Unplanned tissue or organ removal as a result of iAE?: | ◯ Yes<br>◯ No<br>(If unknown, please select "No") |
| iAE Management | |
| Was any surgical repair, medical treatment, or other intervention required?: | ◯ Yes<br>◯ No<br>(If unknown, please select "No". Standard procedural management (e.g. cauterization, use of prothombotic material, small vessel ligation) does not qualify as "repair.") |
| Was there a change in post-operative care due to the iAE?: | ◯ Yes<br>◯ No<br>(If unknown, please select "No") |
| Did the iAE or its management necessitate intensive care admission?: | ◯ Yes<br>◯ No<br>(If unknown, please select "No") |
| Was the intraoperative injury missed, necessitating re-operation within 7 days of index procedure?: | ◯ Yes<br>◯ No<br>(If unknown, please select "No") |
| Bleeding Related | |
| Was blood loss appreciably over normal range for procedure?: | ◯ Yes<br>◯ No<br>(If unknown, please select "No". Per Kazaryan, et al. (2013): "A normal range of blood loss for each particular procedure is subjective in a certain degree, but one can quantify it in regard to different procedures based both on contemporary scientific literature and values typical for own institution") |
| Were 2 or more units of blood products required to manage iAE?: | ◯ Yes<br>◯ No<br>(If unknown, please select "No") |
| iAE Grading | |

| | |
|---|---|
| EAUiaiC | ◯ Grade 0: Event requiring no intervention or change in operative approach, no deviation from planned intraoperative steps<br>◯ Grade 1: Events requiring change in planned intraoperative steps, not life-threatening, no tissue or organ removal. Event address in a controlled manner with no long term side effects<br>◯ Grade 2: Event requiring change in operative approach but NOT life threatening. The event was addressed in a controlled manner, however may have short/long-term side effects<br>◯ Grade 3: Event requiring deviation from planned intraoperative steps, event becoming life threatening but NOT requiring tissue or organ removal<br>◯ Grade 4: Event requiring deviation from planned intraoperative steps and with short/long-term consequences to patient<br>◯ Grade 4A: Requiring tissue or organ removal<br>◯ Grade 4B: Unable to complete planned procedure as planned due to a surgical event or technical issue or unplanned stoma<br>◯ Grade 5A: Wrong site or side open surgery or patient or no consent<br>◯ Grade 5B: Death |
| iAE Severity Classification Scheme<br><br>*Excludes minimally invasive to open conversion | ◯ Class I: Injury requiring no repair within the same procedure (e.g. cauterization, use of prothrombotic material, small vessel ligation)<br>◯ Class II: Injury requiring surgical repair, without organ removal or a change in the originally planned procedure (e.g. any suture repair, patch repair)<br>◯ Class III: Injury requiring tissue or organ removal with completion of the originally planned procedure<br>◯ Class IV: Injury requiring a significant change* and/or incompletion of the originally planned procedure<br>◯ Class V: Missed intraoperative injury requiring re-operation within 7 days<br>◯ Class IV: Intraoperative death |
| iAE Severity Classification Scheme<br><br>Suffix T<br><br>(Add if injury transfusion of ≥2 U blood) | ◯ Yes<br>◯ No |

| | |
|---|---|
| Modified Satava<br><br>†A normal range of blood loss for each particular procedure is subjective in a certain degree, but one can quantify it in regard to different procedures based both on contemporary scientific literature and values typical for own institution | ◯ Grade I: Incidents managed without change of operative approach and without further consequences for the patient. This includes minor injury of adherent or adjacent organs and minimal change of intraoperative tactics and cases with blood loss over normal range†<br>◯ Grade II: Incidents with further consequences for the patient This includes cases requiring limited resection of intraoperatively injured organs or cases with blood loss which is appreciably over normal range†. For laparoscopic/thoracoscopic/endoscopic surgery it includes intraoperative incidents requiring conversion<br>◯ Grade III: Incident leading to significant consequences for patient |
| ClassIntra ® (formerly CLASSIC)<br><br>‡Inclusive of surgery or anesthesia related events | ◯ Grade 0: No deviation from the ideal intraoperative course<br>◯ Grade I: Any deviation from the ideal intraoperative course without the need for any additional treatment or intervention. Patient asymptomatic or with mild symptoms<br>◯ Grade II: Any deviation from the ideal intraoperative course with the need for any additional minor treatment or intervention that is not life threatening and not leading to permanent disability. Patient with moderate symptoms<br>◯ Grade III: Any deviation from the ideal intraoperative course with the need for any additonal moderate or treatment or intervention which is potentially life-threatening and/or potentially leading to permanent disability. Patient with severe symptoms<br>◯ Grade IV: Any deviation from the ideal intraoperative course with the need for any additional major and urgent treatment or intervention. Patient with life threatening symptoms or leading to permanent disability<br>◯ Grade V: Any deviation from the ideal intraoperative course with death of the patient |
| EAES Classification | ◯ Grade 1: Minor error, no damage or corrective action required<br>◯ Grade 2: Minor consequential error requiring correc- tive action but no change in post-operative care<br>◯ Grade 3: Consequential error requiring major correc- tive action and/or change in post-operative pathway<br>◯ Grade 4: Life-threatening complication that requires major or immediate corrective action which led to a significant alteration of the post-operative pathway which may include re-operation or intensive care admission<br>◯ Grade 5: Major consequential error resulting in death |

**SCENARIO 3: Laparoscopic radical prostatectomy. Previous appendicitis with rupture and peritonitis treated with open appendicectomy.**
**Complication/Management: Ascending colon perforation during adhesiolysis, faecal content observed and identification of a relative big perforation. Asked a general surgeon for help. Conversion to open, peritoneal lavage followed by right hemicolectomy and colostomy - finally open radical prostatectomy**

Patient

| | |
|---|---|
| Was the iAE associated with death of the patient?: | ◯ Yes<br>◯ No<br>(If unknown, please select "No") |
| Was the iAE immediately life-threatening?: | ◯ Yes<br>◯ No<br>(If unknown, please select "No") |
| Were there significant consequences to the patient as a result of the iAE?: | ◯ Yes<br>◯ No<br>(If unknown, please select "No". Some examples of iAEs with significant consequences includes injury to or unplanned removal of an otherwise healthy major organ (e.g. unplanned pulmonectomy, nephrectomy) and events that are exceedingly challenging to manage in a controlled manner without potential for long-term patient consequences (e.g. increased risk of postoperative multiorgan failure with major blood transfusion).) |
| Was the incorrect site, side, or surgical approach used without consent?: | ◯ Yes<br>◯ No<br>(If unknown, please select "No") |

Procedure

| | |
|---|---|
| Were there any changes in the ideal intraoperative course related to iAE?: | ◯ Yes<br>◯ No<br>(If unknown, please select "No". Changes in course include minor incidents such as unintended cauterization, equipment malfunction, unanticipated anesthesiologic challenges, or procedural delays regardless of consequence or management.) |
| Was there an unanticipated conversion of approach or significant change to the operative steps of the originally planned procedure due to iAE?: | ◯ Yes<br>◯ No<br>(If unknown, please select "No") |
| Was planned procedure aborted or incomplete due to iAE?: | ◯ Yes<br>◯ No<br>(If unknown, please select "No") |
| Unplanned stoma as a result of iAE?: | ◯ Yes<br>◯ No<br>(If unknown, please select "No") |

| | |
|---|---|
| Unplanned tissue or organ removal as a result of iAE?: | ◯ Yes<br>◯ No<br>(If unknown, please select "No") |
| iAE Management | |
| Was any surgical repair, medical treatment, or other intervention required?: | ◯ Yes<br>◯ No<br>(If unknown, please select "No". Standard procedural management (e.g. cauterization, use of prothombotic material, small vessel ligation) does not qualify as "repair.") |
| Was there a change in post-operative care due to the iAE?: | ◯ Yes<br>◯ No<br>(If unknown, please select "No") |
| Did the iAE or its management necessitate intensive care admission?: | ◯ Yes<br>◯ No<br>(If unknown, please select "No") |
| Was the intraoperative injury missed, necessitating re-operation within 7 days of index procedure?: | ◯ Yes<br>◯ No<br>(If unknown, please select "No") |
| Bleeding Related | |
| Was blood loss appreciably over normal range for procedure?: | ◯ Yes<br>◯ No<br>(If unknown, please select "No". Per Kazaryan, et al. (2013): "A normal range of blood loss for each particular procedure is subjective in a certain degree, but one can quantify it in regard to different procedures based both on contemporary scientific literature and values typical for own institution") |
| Were 2 or more units of blood products required to manage iAE?: | ◯ Yes<br>◯ No<br>(If unknown, please select "No") |
| iAE Grading | |

| | |
|---|---|
| EAUiaiC | ◯ Grade 0: Event requiring no intervention or change in operative approach, no deviation from planned intraoperative steps<br>◯ Grade 1: Events requiring change in planned intraoperative steps, not life-threatening, no tissue or organ removal. Event address in a controlled manner with no long term side effects<br>◯ Grade 2: Event requiring change in operative approach but NOT life threatening. The event was addressed in a controlled manner, however may have short/long-term side effects<br>◯ Grade 3: Event requiring deviation from planned intraoperative steps, event becoming life threatening but NOT requiring tissue or organ removal<br>◯ Grade 4: Event requiring deviation from planned intraoperative steps and with short/long-term consequences to patient<br>◯ Grade 4A: Requiring tissue or organ removal<br>◯ Grade 4B: Unable to complete planned procedure as planned due to a surgical event or technical issue or unplanned stoma<br>◯ Grade 5A: Wrong site or side open surgery or patient or no consent<br>◯ Grade 5B: Death |
| iAE Severity Classification Scheme<br><br>*Excludes minimally invasive to open conversion | ◯ Class I: Injury requiring no repair within the same procedure (e.g. cauterization, use of prothrombotic material, small vessel ligation)<br>◯ Class II: Injury requiring surgical repair, without organ removal or a change in the originally planned procedure (e.g. any suture repair, patch repair)<br>◯ Class III: Injury requiring tissue or organ removal with completion of the originally planned procedure<br>◯ Class IV: Injury requiring a significant change* and/or incompletion of the originally planned procedure<br>◯ Class V: Missed intraoperative injury requiring re-operation within 7 days<br>◯ Class IV: Intraoperative death |
| iAE Severity Classification Scheme<br><br>Suffix T<br><br>(Add if injury transfusion of ≥2 U blood) | ◯ Yes<br>◯ No |

| | |
|---|---|
| Modified Satava<br><br>†A normal range of blood loss for each particular procedure is subjective in a certain degree, but one can quantify it in regard to different procedures based both on contemporary scientific literature and values typical for own institution | ◯ Grade I: Incidents managed without change of operative approach and without further consequences for the patient. This includes minor injury of adherent or adjacent organs and minimal change of intraoperative tactics and cases with blood loss over normal range†<br>◯ Grade II: Incidents with further consequences for the patient This includes cases requiring limited resection of intraoperatively injured organs or cases with blood loss which is appreciably over normal range†. For laparoscopic/thoracoscopic/endoscopic surgery it includes intraoperative incidents requiring conversion<br>◯ Grade III: Incident leading to significant consequences for patient |
| ClassIntra ® (formerly CLASSIC)<br><br>‡Inclusive of surgery or anesthesia related events | ◯ Grade 0: No deviation from the ideal intraoperative course<br>◯ Grade I: Any deviation from the ideal intraoperative course without the need for any additional treatment or intervention. Patient asymptomatic or with mild symptoms<br>◯ Grade II: Any deviation from the ideal intraoperative course with the need for any additional minor treatment or intervention that is not life threatening and not leading to permanent disability. Patient with moderate symptoms<br>◯ Grade III: Any deviation from the ideal intraoperative course with the need for any additonal moderate or treatment or intervention which is potentially life-threatening and/or potentially leading to permanent disability. Patient with severe symptoms<br>◯ Grade IV: Any deviation from the ideal intraoperative course with the need for any additional major and urgent treatment or intervention. Patient with life threatening symptoms or leading to permanent disability<br>◯ Grade V: Any deviation from the ideal intraoperative course with death of the patient |
| EAES Classification | ◯ Grade 1: Minor error, no damage or corrective action required<br>◯ Grade 2: Minor consequential error requiring correc- tive action but no change in post-operative care<br>◯ Grade 3: Consequential error requiring major correc- tive action and/or change in post-operative pathway<br>◯ Grade 4: Life-threatening complication that requires major or immediate corrective action which led to a significant alteration of the post-operative pathway which may include re-operation or intensive care admission<br>◯ Grade 5: Major consequential error resulting in death |

#### SCENARIO 4:

#### Injury of renal artery resulting in necessity to remove a healthy kidney during adrenalectomy

Patient

| | |
|---|---|
| Was the iAE associated with death of the patient?: | ◯ Yes<br>◯ No<br>(If unknown, please select "No") |
| Was the iAE immediately life-threatening?: | ◯ Yes<br>◯ No<br>(If unknown, please select "No") |
| Were there significant consequences to the patient as a result of the iAE?: | ◯ Yes<br>◯ No<br>(If unknown, please select "No". Some examples of iAEs with significant consequences includes injury to or unplanned removal of an otherwise healthy major organ (e.g. unplanned pulmonectomy, nephrectomy) and events that are exceedingly challenging to manage in a controlled manner without potential for long-term patient consequences (e.g. increased risk of postoperative multiorgan failure with major blood transfusion).) |
| Was the incorrect site, side, or surgical approach used without consent?: | ◯ Yes<br>◯ No<br>(If unknown, please select "No") |

Procedure

| | |
|---|---|
| Were there any changes in the ideal intraoperative course related to iAE?: | ◯ Yes<br>◯ No<br>(If unknown, please select "No". Changes in course include minor incidents such as unintended cauterization, equipment malfunction, unanticipated anesthesiologic challenges, or procedural delays regardless of consequence or management.) |
| Was there an unanticipated conversion of approach or significant change to the operative steps of the originally planned procedure due to iAE?: | ◯ Yes<br>◯ No<br>(If unknown, please select "No") |
| Was planned procedure aborted or incomplete due to iAE?: | ◯ Yes<br>◯ No<br>(If unknown, please select "No") |
| Unplanned stoma as a result of iAE?: | ◯ Yes<br>◯ No<br>(If unknown, please select "No") |
| Unplanned tissue or organ removal as a result of iAE?: | ◯ Yes<br>◯ No<br>(If unknown, please select "No") |

iAE Management

| | |
|---|---|
| Was any surgical repair, medical treatment, or other intervention required?: | ◯ Yes<br>◯ No<br>(If unknown, please select "No". Standard procedural management (e.g. cauterization, use of prothombotic material, small vessel ligation) does not qualify as "repair.") |
| Was there a change in post-operative care due to the iAE?: | ◯ Yes<br>◯ No<br>(If unknown, please select "No") |
| Did the iAE or its management necessitate intensive care admission?: | ◯ Yes<br>◯ No<br>(If unknown, please select "No") |
| Was the intraoperative injury missed, necessitating re-operation within 7 days of index procedure?: | ◯ Yes<br>◯ No<br>(If unknown, please select "No") |
| Bleeding Related | |
| Was blood loss appreciably over normal range for procedure?: | ◯ Yes<br>◯ No<br>(If unknown, please select "No". Per Kazaryan, et al. (2013): "A normal range of blood loss for each particular procedure is subjective in a certain degree, but one can quantify it in regard to different procedures based both on contemporary scientific literature and values typical for own institution") |
| Were 2 or more units of blood products required to manage iAE?: | ◯ Yes<br>◯ No<br>(If unknown, please select "No") |
| iAE Grading | |

| | |
|---|---|
| EAUiaiC | ◯ Grade 0: Event requiring no intervention or change in operative approach, no deviation from planned intraoperative steps<br>◯ Grade 1: Events requiring change in planned intraoperative steps, not life-threatening, no tissue or organ removal. Event address in a controlled manner with no long term side effects<br>◯ Grade 2: Event requiring change in operative approach but NOT life threatening. The event was addressed in a controlled manner, however may have short/long-term side effects<br>◯ Grade 3: Event requiring deviation from planned intraoperative steps, event becoming life threatening but NOT requiring tissue or organ removal<br>◯ Grade 4: Event requiring deviation from planned intraoperative steps and with short/long-term consequences to patient<br>◯ Grade 4A: Requiring tissue or organ removal<br>◯ Grade 4B: Unable to complete planned procedure as planned due to a surgical event or technical issue or unplanned stoma<br>◯ Grade 5A: Wrong site or side open surgery or patient or no consent<br>◯ Grade 5B: Death |
| iAE Severity Classification Scheme<br><br>*Excludes minimally invasive to open conversion | ◯ Class I: Injury requiring no repair within the same procedure (e.g. cauterization, use of prothrombotic material, small vessel ligation)<br>◯ Class II: Injury requiring surgical repair, without organ removal or a change in the originally planned procedure (e.g. any suture repair, patch repair)<br>◯ Class III: Injury requiring tissue or organ removal with completion of the originally planned procedure<br>◯ Class IV: Injury requiring a significant change* and/or incompletion of the originally planned procedure<br>◯ Class V: Missed intraoperative injury requiring re-operation within 7 days<br>◯ Class IV: Intraoperative death |
| iAE Severity Classification Scheme<br><br>Suffix T<br><br>(Add if injury transfusion of ≥2 U blood) | ◯ Yes<br>◯ No |

| | |
|---|---|
| Modified Satava<br><br>†A normal range of blood loss for each particular procedure is subjective in a certain degree, but one can quantify it in regard to different procedures based both on contemporary scientific literature and values typical for own institution | ◯ Grade I: Incidents managed without change of operative approach and without further consequences for the patient. This includes minor injury of adherent or adjacent organs and minimal change of intraoperative tactics and cases with blood loss over normal range†<br>◯ Grade II: Incidents with further consequences for the patient This includes cases requiring limited resection of intraoperatively injured organs or cases with blood loss which is appreciably over normal range†. For laparoscopic/thoracoscopic/endoscopic surgery it includes intraoperative incidents requiring conversion<br>◯ Grade III: Incident leading to significant consequences for patient |
| ClassIntra ® (formerly CLASSIC)<br><br>‡Inclusive of surgery or anesthesia related events | ◯ Grade 0: No deviation from the ideal intraoperative course<br>◯ Grade I: Any deviation from the ideal intraoperative course without the need for any additional treatment or intervention. Patient asymptomatic or with mild symptoms<br>◯ Grade II: Any deviation from the ideal intraoperative course with the need for any additional minor treatment or intervention that is not life threatening and not leading to permanent disability. Patient with moderate symptoms<br>◯ Grade III: Any deviation from the ideal intraoperative course with the need for any additonal moderate or treatment or intervention which is potentially life-threatening and/or potentially leading to permanent disability. Patient with severe symptoms<br>◯ Grade IV: Any deviation from the ideal intraoperative course with the need for any additional major and urgent treatment or intervention. Patient with life threatening symptoms or leading to permanent disability<br>◯ Grade V: Any deviation from the ideal intraoperative course with death of the patient |
| EAES Classification | ◯ Grade 1: Minor error, no damage or corrective action required<br>◯ Grade 2: Minor consequential error requiring correc- tive action but no change in post-operative care<br>◯ Grade 3: Consequential error requiring major correc- tive action and/or change in post-operative pathway<br>◯ Grade 4: Life-threatening complication that requires major or immediate corrective action which led to a significant alteration of the post-operative pathway which may include re-operation or intensive care admission<br>◯ Grade 5: Major consequential error resulting in death |

**SCENARIO 5:**

**"Right laparoscopic nephrectomy.**
**Complication/Management: Unable to disengage endo GIA after stapling the renal vein.**
**Converted to open to complete the procedure "**

Patient

| | |
|---|---|
| Was the iAE associated with death of the patient?: | ◯ Yes<br>◯ No<br>(If unknown, please select "No") |
| Was the iAE immediately life-threatening?: | ◯ Yes<br>◯ No<br>(If unknown, please select "No") |
| Were there significant consequences to the patient as a result of the iAE?: | ◯ Yes<br>◯ No<br>(If unknown, please select "No". Some examples of iAEs with significant consequences includes injury to or unplanned removal of an otherwise healthy major organ (e.g. unplanned pulmonectomy, nephrectomy) and events that are exceedingly challenging to manage in a controlled manner without potential for long-term patient consequences (e.g. increased risk of postoperative multiorgan failure with major blood transfusion).) |
| Was the incorrect site, side, or surgical approach used without consent?: | ◯ Yes<br>◯ No<br>(If unknown, please select "No") |

Procedure

| | |
|---|---|
| Were there any changes in the ideal intraoperative course related to iAE?: | ◯ Yes<br>◯ No<br>(If unknown, please select "No". Changes in course include minor incidents such as unintended cauterization, equipment malfunction, unanticipated anesthesiologic challenges, or procedural delays regardless of consequence or management.) |
| Was there an unanticipated conversion of approach or significant change to the operative steps of the originally planned procedure due to iAE?: | ◯ Yes<br>◯ No<br>(If unknown, please select "No") |
| Was planned procedure aborted or incomplete due to iAE?: | ◯ Yes<br>◯ No<br>(If unknown, please select "No") |
| Unplanned stoma as a result of iAE?: | ◯ Yes<br>◯ No<br>(If unknown, please select "No") |
| Unplanned tissue or organ removal as a result of iAE?: | ◯ Yes<br>◯ No<br>(If unknown, please select "No") |

| | |
|---|---|
| iAE Management | |
| Was any surgical repair, medical treatment, or other intervention required?: | ◯ Yes<br>◯ No<br>(If unknown, please select "No". Standard procedural management (e.g. cauterization, use of prothombotic material, small vessel ligation) does not qualify as "repair.") |
| Was there a change in post-operative care due to the iAE?: | ◯ Yes<br>◯ No<br>(If unknown, please select "No") |
| Did the iAE or its management necessitate intensive care admission?: | ◯ Yes<br>◯ No<br>(If unknown, please select "No") |
| Was the intraoperative injury missed, necessitating re-operation within 7 days of index procedure?: | ◯ Yes<br>◯ No<br>(If unknown, please select "No") |
| Bleeding Related | |
| Was blood loss appreciably over normal range for procedure?: | ◯ Yes<br>◯ No<br>(If unknown, please select "No". Per Kazaryan, et al. (2013): "A normal range of blood loss for each particular procedure is subjective in a certain degree, but one can quantify it in regard to different procedures based both on contemporary scientific literature and values typical for own institution") |
| Were 2 or more units of blood products required to manage iAE?: | ◯ Yes<br>◯ No<br>(If unknown, please select "No") |
| iAE Grading | |

| | |
|---|---|
| EAUiaiC | ◯ Grade 0: Event requiring no intervention or change in operative approach, no deviation from planned intraoperative steps<br>◯ Grade 1: Events requiring change in planned intraoperative steps, not life-threatening, no tissue or organ removal. Event address in a controlled manner with no long term side effects<br>◯ Grade 2: Event requiring change in operative approach but NOT life threatening. The event was addressed in a controlled manner, however may have short/long-term side effects<br>◯ Grade 3: Event requiring deviation from planned intraoperative steps, event becoming life threatening but NOT requiring tissue or organ removal<br>◯ Grade 4: Event requiring deviation from planned intraoperative steps and with short/long-term consequences to patient<br>◯ Grade 4A: Requiring tissue or organ removal<br>◯ Grade 4B: Unable to complete planned procedure as planned due to a surgical event or technical issue or unplanned stoma<br>◯ Grade 5A: Wrong site or side open surgery or patient or no consent<br>◯ Grade 5B: Death |
| iAE Severity Classification Scheme<br><br>*Excludes minimally invasive to open conversion | ◯ Class I: Injury requiring no repair within the same procedure (e.g. cauterization, use of prothrombotic material, small vessel ligation)<br>◯ Class II: Injury requiring surgical repair, without organ removal or a change in the originally planned procedure (e.g. any suture repair, patch repair)<br>◯ Class III: Injury requiring tissue or organ removal with completion of the originally planned procedure<br>◯ Class IV: Injury requiring a significant change* and/or incompletion of the originally planned procedure<br>◯ Class V: Missed intraoperative injury requiring re-operation within 7 days<br>◯ Class IV: Intraoperative death |
| iAE Severity Classification Scheme<br><br>Suffix T<br><br>(Add if injury transfusion of ≥2 U blood) | ◯ Yes<br>◯ No |

| | |
|---|---|
| Modified Satava<br><br>†A normal range of blood loss for each particular procedure is subjective in a certain degree, but one can quantify it in regard to different procedures based both on contemporary scientific literature and values typical for own institution | ◯ Grade I: Incidents managed without change of operative approach and without further consequences for the patient. This includes minor injury of adherent or adjacent organs and minimal change of intraoperative tactics and cases with blood loss over normal range†<br>◯ Grade II: Incidents with further consequences for the patient This includes cases requiring limited resection of intraoperatively injured organs or cases with blood loss which is appreciably over normal range†. For laparoscopic/thoracoscopic/endoscopic surgery it includes intraoperative incidents requiring conversion<br>◯ Grade III: Incident leading to significant consequences for patient |
| ClassIntra ® (formerly CLASSIC)<br><br>‡Inclusive of surgery or anesthesia related events | ◯ Grade 0: No deviation from the ideal intraoperative course<br>◯ Grade I: Any deviation from the ideal intraoperative course without the need for any additional treatment or intervention. Patient asymptomatic or with mild symptoms<br>◯ Grade II: Any deviation from the ideal intraoperative course with the need for any additional minor treatment or intervention that is not life threatening and not leading to permanent disability. Patient with moderate symptoms<br>◯ Grade III: Any deviation from the ideal intraoperative course with the need for any additonal moderate or treatment or intervention which is potentially life-threatening and/or potentially leading to permanent disability. Patient with severe symptoms<br>◯ Grade IV: Any deviation from the ideal intraoperative course with the need for any additional major and urgent treatment or intervention. Patient with life threatening symptoms or leading to permanent disability<br>◯ Grade V: Any deviation from the ideal intraoperative course with death of the patient |
| EAES Classification | ◯ Grade 1: Minor error, no damage or corrective action required<br>◯ Grade 2: Minor consequential error requiring correc- tive action but no change in post-operative care<br>◯ Grade 3: Consequential error requiring major correc- tive action and/or change in post-operative pathway<br>◯ Grade 4: Life-threatening complication that requires major or immediate corrective action which led to a significant alteration of the post-operative pathway which may include re-operation or intensive care admission<br>◯ Grade 5: Major consequential error resulting in death |

**SCENARIO 6:**

**Right robotic pyeloplasty. Previous right endopyelotomy**
**Complication/Management: Crossing large renal vein injury. Repair using a 5 – 0 prolene.**
**Completed procedure**

Patient

| | |
|---|---|
| Was the iAE associated with death of the patient?: | ◯ Yes<br>◯ No<br>(If unknown, please select "No") |
| Was the iAE immediately life-threatening?: | ◯ Yes<br>◯ No<br>(If unknown, please select "No") |
| Were there significant consequences to the patient as a result of the iAE?: | ◯ Yes<br>◯ No<br>(If unknown, please select "No". Some examples of iAEs with significant consequences includes injury to or unplanned removal of an otherwise healthy major organ (e.g. unplanned pulmonectomy, nephrectomy) and events that are exceedingly challenging to manage in a controlled manner without potential for long-term patient consequences (e.g. increased risk of postoperative multiorgan failure with major blood transfusion).) |
| Was the incorrect site, side, or surgical approach used without consent?: | ◯ Yes<br>◯ No<br>(If unknown, please select "No") |

Procedure

| | |
|---|---|
| Were there any changes in the ideal intraoperative course related to iAE?: | ◯ Yes<br>◯ No<br>(If unknown, please select "No". Changes in course include minor incidents such as unintended cauterization, equipment malfunction, unanticipated anesthesiologic challenges, or procedural delays regardless of consequence or management.) |
| Was there an unanticipated conversion of approach or significant change to the operative steps of the originally planned procedure due to iAE?: | ◯ Yes<br>◯ No<br>(If unknown, please select "No") |
| Was planned procedure aborted or incomplete due to iAE?: | ◯ Yes<br>◯ No<br>(If unknown, please select "No") |
| Unplanned stoma as a result of iAE?: | ◯ Yes<br>◯ No<br>(If unknown, please select "No") |
| Unplanned tissue or organ removal as a result of iAE?: | ◯ Yes<br>◯ No<br>(If unknown, please select "No") |

iAE Management

| | |
|---|---|
| Was any surgical repair, medical treatment, or other intervention required?: | ◯ Yes<br>◯ No<br>(If unknown, please select "No". Standard procedural management (e.g. cauterization, use of prothombotic material, small vessel ligation) does not qualify as "repair.") |
| Was there a change in post-operative care due to the iAE?: | ◯ Yes<br>◯ No<br>(If unknown, please select "No") |
| Did the iAE or its management necessitate intensive care admission?: | ◯ Yes<br>◯ No<br>(If unknown, please select "No") |
| Was the intraoperative injury missed, necessitating re-operation within 7 days of index procedure?: | ◯ Yes<br>◯ No<br>(If unknown, please select "No") |

Bleeding Related

| | |
|---|---|
| Was blood loss appreciably over normal range for procedure?: | ◯ Yes<br>◯ No<br>(If unknown, please select "No". Per Kazaryan, et al. (2013): "A normal range of blood loss for each particular procedure is subjective in a certain degree, but one can quantify it in regard to different procedures based both on contemporary scientific literature and values typical for own institution") |
| Were 2 or more units of blood products required to manage iAE?: | ◯ Yes<br>◯ No<br>(If unknown, please select "No") |

iAE Grading

| | |
|---|---|
| EAUiaiC | ◯ Grade 0: Event requiring no intervention or change in operative approach, no deviation from planned intraoperative steps<br>◯ Grade 1: Events requiring change in planned intraoperative steps, not life-threatening, no tissue or organ removal. Event address in a controlled manner with no long term side effects<br>◯ Grade 2: Event requiring change in operative approach but NOT life threatening. The event was addressed in a controlled manner, however may have short/long-term side effects<br>◯ Grade 3: Event requiring deviation from planned intraoperative steps, event becoming life threatening but NOT requiring tissue or organ removal<br>◯ Grade 4: Event requiring deviation from planned intraoperative steps and with short/long-term consequences to patient<br>◯ Grade 4A: Requiring tissue or organ removal<br>◯ Grade 4B: Unable to complete planned procedure as planned due to a surgical event or technical issue or unplanned stoma<br>◯ Grade 5A: Wrong site or side open surgery or patient or no consent<br>◯ Grade 5B: Death |
| iAE Severity Classification Scheme<br><br>*Excludes minimally invasive to open conversion | ◯ Class I: Injury requiring no repair within the same procedure (e.g. cauterization, use of prothrombotic material, small vessel ligation)<br>◯ Class II: Injury requiring surgical repair, without organ removal or a change in the originally planned procedure (e.g. any suture repair, patch repair)<br>◯ Class III: Injury requiring tissue or organ removal with completion of the originally planned procedure<br>◯ Class IV: Injury requiring a significant change* and/or incompletion of the originally planned procedure<br>◯ Class V: Missed intraoperative injury requiring re-operation within 7 days<br>◯ Class IV: Intraoperative death |
| iAE Severity Classification Scheme<br><br>Suffix T<br><br>(Add if injury transfusion of ≥2 U blood) | ◯ Yes<br>◯ No |

| | |
|---|---|
| Modified Satava<br><br>†A normal range of blood loss for each particular procedure is subjective in a certain degree, but one can quantify it in regard to different procedures based both on contemporary scientific literature and values typical for own institution | ◯ Grade I: Incidents managed without change of operative approach and without further consequences for the patient. This includes minor injury of adherent or adjacent organs and minimal change of intraoperative tactics and cases with blood loss over normal range†<br>◯ Grade II: Incidents with further consequences for the patient This includes cases requiring limited resection of intraoperatively injured organs or cases with blood loss which is appreciably over normal range†. For laparoscopic/thoracoscopic/endoscopic surgery it includes intraoperative incidents requiring conversion<br>◯ Grade III: Incident leading to significant consequences for patient |
| ClassIntra ® (formerly CLASSIC)<br><br>‡Inclusive of surgery or anesthesia related events | ◯ Grade 0: No deviation from the ideal intraoperative course<br>◯ Grade I: Any deviation from the ideal intraoperative course without the need for any additional treatment or intervention. Patient asymptomatic or with mild symptoms<br>◯ Grade II: Any deviation from the ideal intraoperative course with the need for any additional minor treatment or intervention that is not life threatening and not leading to permanent disability. Patient with moderate symptoms<br>◯ Grade III: Any deviation from the ideal intraoperative course with the need for any additonal moderate or treatment or intervention which is potentially life-threatening and/or potentially leading to permanent disability. Patient with severe symptoms<br>◯ Grade IV: Any deviation from the ideal intraoperative course with the need for any additional major and urgent treatment or intervention. Patient with life threatening symptoms or leading to permanent disability<br>◯ Grade V: Any deviation from the ideal intraoperative course with death of the patient |
| EAES Classification | ◯ Grade 1: Minor error, no damage or corrective action required<br>◯ Grade 2: Minor consequential error requiring correc- tive action but no change in post-operative care<br>◯ Grade 3: Consequential error requiring major correc- tive action and/or change in post-operative pathway<br>◯ Grade 4: Life-threatening complication that requires major or immediate corrective action which led to a significant alteration of the post-operative pathway which may include re-operation or intensive care admission<br>◯ Grade 5: Major consequential error resulting in death |

**SCENARIO 7:**

**Insertion of sacral nerve stimulator on the right side. Previous intravesical botox Complication/Management: Stimulator was inserted on the Left side. Mistake was noted in theatre and procedure was done on the RIGHT side. Mistake corrected**

Patient

| | |
|---|---|
| Was the iAE associated with death of the patient?: | ◯ Yes<br>◯ No<br>(If unknown, please select "No") |
| Was the iAE immediately life-threatening?: | ◯ Yes<br>◯ No<br>(If unknown, please select "No") |
| Were there significant consequences to the patient as a result of the iAE?: | ◯ Yes<br>◯ No<br>(If unknown, please select "No". Some examples of iAEs with significant consequences includes injury to or unplanned removal of an otherwise healthy major organ (e.g. unplanned pulmonectomy, nephrectomy) and events that are exceedingly challenging to manage in a controlled manner without potential for long-term patient consequences (e.g. increased risk of postoperative multiorgan failure with major blood transfusion).) |
| Was the incorrect site, side, or surgical approach used without consent?: | ◯ Yes<br>◯ No<br>(If unknown, please select "No") |

Procedure

| | |
|---|---|
| Were there any changes in the ideal intraoperative course related to iAE?: | ◯ Yes<br>◯ No<br>(If unknown, please select "No". Changes in course include minor incidents such as unintended cauterization, equipment malfunction, unanticipated anesthesiologic challenges, or procedural delays regardless of consequence or management.) |
| Was there an unanticipated conversion of approach or significant change to the operative steps of the originally planned procedure due to iAE?: | ◯ Yes<br>◯ No<br>(If unknown, please select "No") |
| Was planned procedure aborted or incomplete due to iAE?: | ◯ Yes<br>◯ No<br>(If unknown, please select "No") |
| Unplanned stoma as a result of iAE?: | ◯ Yes<br>◯ No<br>(If unknown, please select "No") |
| Unplanned tissue or organ removal as a result of iAE?: | ◯ Yes<br>◯ No<br>(If unknown, please select "No") |

iAE Management

| | |
|---|---|
| Was any surgical repair, medical treatment, or other intervention required?: | ◯ Yes<br>◯ No<br>(If unknown, please select "No". Standard procedural management (e.g. cauterization, use of prothombotic material, small vessel ligation) does not qualify as "repair.") |
| Was there a change in post-operative care due to the iAE?: | ◯ Yes<br>◯ No<br>(If unknown, please select "No") |
| Did the iAE or its management necessitate intensive care admission?: | ◯ Yes<br>◯ No<br>(If unknown, please select "No") |
| Was the intraoperative injury missed, necessitating re-operation within 7 days of index procedure?: | ◯ Yes<br>◯ No<br>(If unknown, please select "No") |

Bleeding Related

| | |
|---|---|
| Was blood loss appreciably over normal range for procedure?: | ◯ Yes<br>◯ No<br>(If unknown, please select "No". Per Kazaryan, et al. (2013): "A normal range of blood loss for each particular procedure is subjective in a certain degree, but one can quantify it in regard to different procedures based both on contemporary scientific literature and values typical for own institution") |
| Were 2 or more units of blood products required to manage iAE?: | ◯ Yes<br>◯ No<br>(If unknown, please select "No") |

iAE Grading

| | |
|---|---|
| EAUiaiC | ◯ Grade 0: Event requiring no intervention or change in operative approach, no deviation from planned intraoperative steps<br>◯ Grade 1: Events requiring change in planned intraoperative steps, not life-threatening, no tissue or organ removal. Event address in a controlled manner with no long term side effects<br>◯ Grade 2: Event requiring change in operative approach but NOT life threatening. The event was addressed in a controlled manner, however may have short/long-term side effects<br>◯ Grade 3: Event requiring deviation from planned intraoperative steps, event becoming life threatening but NOT requiring tissue or organ removal<br>◯ Grade 4: Event requiring deviation from planned intraoperative steps and with short/long-term consequences to patient<br>◯ Grade 4A: Requiring tissue or organ removal<br>◯ Grade 4B: Unable to complete planned procedure as planned due to a surgical event or technical issue or unplanned stoma<br>◯ Grade 5A: Wrong site or side open surgery or patient or no consent<br>◯ Grade 5B: Death |
| iAE Severity Classification Scheme<br><br>*Excludes minimally invasive to open conversion | ◯ Class I: Injury requiring no repair within the same procedure (e.g. cauterization, use of prothrombotic material, small vessel ligation)<br>◯ Class II: Injury requiring surgical repair, without organ removal or a change in the originally planned procedure (e.g. any suture repair, patch repair)<br>◯ Class III: Injury requiring tissue or organ removal with completion of the originally planned procedure<br>◯ Class IV: Injury requiring a significant change* and/or incompletion of the originally planned procedure<br>◯ Class V: Missed intraoperative injury requiring re-operation within 7 days<br>◯ Class IV: Intraoperative death |
| iAE Severity Classification Scheme<br><br>Suffix T<br><br>(Add if injury transfusion of ≥2 U blood) | ◯ Yes<br>◯ No |

| | |
|---|---|
| Modified Satava<br><br>†A normal range of blood loss for each particular procedure is subjective in a certain degree, but one can quantify it in regard to different procedures based both on contemporary scientific literature and values typical for own institution | ◯ Grade I: Incidents managed without change of operative approach and without further consequences for the patient. This includes minor injury of adherent or adjacent organs and minimal change of intraoperative tactics and cases with blood loss over normal range†<br>◯ Grade II: Incidents with further consequences for the patient This includes cases requiring limited resection of intraoperatively injured organs or cases with blood loss which is appreciably over normal range†. For laparoscopic/thoracoscopic/endoscopic surgery it includes intraoperative incidents requiring conversion<br>◯ Grade III: Incident leading to significant consequences for patient |
| ClassIntra ® (formerly CLASSIC)<br><br>‡Inclusive of surgery or anesthesia related events | ◯ Grade 0: No deviation from the ideal intraoperative course<br>◯ Grade I: Any deviation from the ideal intraoperative course without the need for any additional treatment or intervention. Patient asymptomatic or with mild symptoms<br>◯ Grade II: Any deviation from the ideal intraoperative course with the need for any additional minor treatment or intervention that is not life threatening and not leading to permanent disability. Patient with moderate symptoms<br>◯ Grade III: Any deviation from the ideal intraoperative course with the need for any additonal moderate or treatment or intervention which is potentially life-threatening and/or potentially leading to permanent disability. Patient with severe symptoms<br>◯ Grade IV: Any deviation from the ideal intraoperative course with the need for any additional major and urgent treatment or intervention. Patient with life threatening symptoms or leading to permanent disability<br>◯ Grade V: Any deviation from the ideal intraoperative course with death of the patient |
| EAES Classification | ◯ Grade 1: Minor error, no damage or corrective action required<br>◯ Grade 2: Minor consequential error requiring correc- tive action but no change in post-operative care<br>◯ Grade 3: Consequential error requiring major correc- tive action and/or change in post-operative pathway<br>◯ Grade 4: Life-threatening complication that requires major or immediate corrective action which led to a significant alteration of the post-operative pathway which may include re-operation or intensive care admission<br>◯ Grade 5: Major consequential error resulting in death |

#### SCENARIO 8:

#### Conversion to conventional laparotomy to manage intraoperative incident

| | |
|---|---|
| Patient | |
| Was the iAE associated with death of the patient?: | ◯ Yes<br>◯ No<br>(If unknown, please select "No") |
| Was the iAE immediately life-threatening?: | ◯ Yes<br>◯ No<br>(If unknown, please select "No") |
| Were there significant consequences to the patient as a result of the iAE?: | ◯ Yes<br>◯ No<br>(If unknown, please select "No". Some examples of iAEs with significant consequences includes injury to or unplanned removal of an otherwise healthy major organ (e.g. unplanned pulmonectomy, nephrectomy) and events that are exceedingly challenging to manage in a controlled manner without potential for long-term patient consequences (e.g. increased risk of postoperative multiorgan failure with major blood transfusion).) |
| Was the incorrect site, side, or surgical approach used without consent?: | ◯ Yes<br>◯ No<br>(If unknown, please select "No") |
| Procedure | |
| Were there any changes in the ideal intraoperative course related to iAE?: | ◯ Yes<br>◯ No<br>(If unknown, please select "No". Changes in course include minor incidents such as unintended cauterization, equipment malfunction, unanticipated anesthesiologic challenges, or procedural delays regardless of consequence or management.) |
| Was there an unanticipated conversion of approach or significant change to the operative steps of the originally planned procedure due to iAE?: | ◯ Yes<br>◯ No<br>(If unknown, please select "No") |
| Was planned procedure aborted or incomplete due to iAE?: | ◯ Yes<br>◯ No<br>(If unknown, please select "No") |
| Unplanned stoma as a result of iAE?: | ◯ Yes<br>◯ No<br>(If unknown, please select "No") |
| Unplanned tissue or organ removal as a result of iAE?: | ◯ Yes<br>◯ No<br>(If unknown, please select "No") |
| iAE Management | |

| | |
|---|---|
| Was any surgical repair, medical treatment, or other intervention required?: | ◯ Yes<br>◯ No<br>(If unknown, please select "No". Standard procedural management (e.g. cauterization, use of prothombotic material, small vessel ligation) does not qualify as "repair.") |
| Was there a change in post-operative care due to the iAE?: | ◯ Yes<br>◯ No<br>(If unknown, please select "No") |
| Did the iAE or its management necessitate intensive care admission?: | ◯ Yes<br>◯ No<br>(If unknown, please select "No") |
| Was the intraoperative injury missed, necessitating re-operation within 7 days of index procedure?: | ◯ Yes<br>◯ No<br>(If unknown, please select "No") |
| Bleeding Related | |
| Was blood loss appreciably over normal range for procedure?: | ◯ Yes<br>◯ No<br>(If unknown, please select "No". Per Kazaryan, et al. (2013): "A normal range of blood loss for each particular procedure is subjective in a certain degree, but one can quantify it in regard to different procedures based both on contemporary scientific literature and values typical for own institution") |
| Were 2 or more units of blood products required to manage iAE?: | ◯ Yes<br>◯ No<br>(If unknown, please select "No") |
| iAE Grading | |

| | |
|---|---|
| EAUiaiC | ◯ Grade 0: Event requiring no intervention or change in operative approach, no deviation from planned intraoperative steps<br>◯ Grade 1: Events requiring change in planned intraoperative steps, not life-threatening, no tissue or organ removal. Event address in a controlled manner with no long term side effects<br>◯ Grade 2: Event requiring change in operative approach but NOT life threatening. The event was addressed in a controlled manner, however may have short/long-term side effects<br>◯ Grade 3: Event requiring deviation from planned intraoperative steps, event becoming life threatening but NOT requiring tissue or organ removal<br>◯ Grade 4: Event requiring deviation from planned intraoperative steps and with short/long-term consequences to patient<br>◯ Grade 4A: Requiring tissue or organ removal<br>◯ Grade 4B: Unable to complete planned procedure as planned due to a surgical event or technical issue or unplanned stoma<br>◯ Grade 5A: Wrong site or side open surgery or patient or no consent<br>◯ Grade 5B: Death |
| iAE Severity Classification Scheme<br><br>*Excludes minimally invasive to open conversion | ◯ Class I: Injury requiring no repair within the same procedure (e.g. cauterization, use of prothrombotic material, small vessel ligation)<br>◯ Class II: Injury requiring surgical repair, without organ removal or a change in the originally planned procedure (e.g. any suture repair, patch repair)<br>◯ Class III: Injury requiring tissue or organ removal with completion of the originally planned procedure<br>◯ Class IV: Injury requiring a significant change* and/or incompletion of the originally planned procedure<br>◯ Class V: Missed intraoperative injury requiring re-operation within 7 days<br>◯ Class IV: Intraoperative death |
| iAE Severity Classification Scheme<br><br>Suffix T<br><br>(Add if injury transfusion of ≥2 U blood) | ◯ Yes<br>◯ No |

| | |
|---|---|
| Modified Satava<br><br>†A normal range of blood loss for each particular procedure is subjective in a certain degree, but one can quantify it in regard to different procedures based both on contemporary scientific literature and values typical for own institution | ◯ Grade I: Incidents managed without change of operative approach and without further consequences for the patient. This includes minor injury of adherent or adjacent organs and minimal change of intraoperative tactics and cases with blood loss over normal range†<br>◯ Grade II: Incidents with further consequences for the patient This includes cases requiring limited resection of intraoperatively injured organs or cases with blood loss which is appreciably over normal range†. For laparoscopic/thoracoscopic/endoscopic surgery it includes intraoperative incidents requiring conversion<br>◯ Grade III: Incident leading to significant consequences for patient |
| ClassIntra ® (formerly CLASSIC)<br><br>‡Inclusive of surgery or anesthesia related events | ◯ Grade 0: No deviation from the ideal intraoperative course<br>◯ Grade I: Any deviation from the ideal intraoperative course without the need for any additional treatment or intervention. Patient asymptomatic or with mild symptoms<br>◯ Grade II: Any deviation from the ideal intraoperative course with the need for any additional minor treatment or intervention that is not life threatening and not leading to permanent disability. Patient with moderate symptoms<br>◯ Grade III: Any deviation from the ideal intraoperative course with the need for any additonal moderate or treatment or intervention which is potentially life-threatening and/or potentially leading to permanent disability. Patient with severe symptoms<br>◯ Grade IV: Any deviation from the ideal intraoperative course with the need for any additional major and urgent treatment or intervention. Patient with life threatening symptoms or leading to permanent disability<br>◯ Grade V: Any deviation from the ideal intraoperative course with death of the patient |
| EAES Classification | ◯ Grade 1: Minor error, no damage or corrective action required<br>◯ Grade 2: Minor consequential error requiring correc- tive action but no change in post-operative care<br>◯ Grade 3: Consequential error requiring major correc- tive action and/or change in post-operative pathway<br>◯ Grade 4: Life-threatening complication that requires major or immediate corrective action which led to a significant alteration of the post-operative pathway which may include re-operation or intensive care admission<br>◯ Grade 5: Major consequential error resulting in death |

#### SCENARIO 9:

#### Splenic injury requiring splenectomy

Patient

| | |
|---|---|
| Was the iAE associated with death of the patient?: | ◯ Yes<br>◯ No<br>(If unknown, please select "No") |
| Was the iAE immediately life-threatening?: | ◯ Yes<br>◯ No<br>(If unknown, please select "No") |
| Were there significant consequences to the patient as a result of the iAE?: | ◯ Yes<br>◯ No<br>(If unknown, please select "No". Some examples of iAEs with significant consequences includes injury to or unplanned removal of an otherwise healthy major organ (e.g. unplanned pulmonectomy, nephrectomy) and events that are exceedingly challenging to manage in a controlled manner without potential for long-term patient consequences (e.g. increased risk of postoperative multiorgan failure with major blood transfusion).) |
| Was the incorrect site, side, or surgical approach used without consent?: | ◯ Yes<br>◯ No<br>(If unknown, please select "No") |

Procedure

| | |
|---|---|
| Were there any changes in the ideal intraoperative course related to iAE?: | ◯ Yes<br>◯ No<br>(If unknown, please select "No". Changes in course include minor incidents such as unintended cauterization, equipment malfunction, unanticipated anesthesiologic challenges, or procedural delays regardless of consequence or management.) |
| Was there an unanticipated conversion of approach or significant change to the operative steps of the originally planned procedure due to iAE?: | ◯ Yes<br>◯ No<br>(If unknown, please select "No") |
| Was planned procedure aborted or incomplete due to iAE?: | ◯ Yes<br>◯ No<br>(If unknown, please select "No") |
| Unplanned stoma as a result of iAE?: | ◯ Yes<br>◯ No<br>(If unknown, please select "No") |
| Unplanned tissue or organ removal as a result of iAE?: | ◯ Yes<br>◯ No<br>(If unknown, please select "No") |

iAE Management

| | |
|---|---|
| Was any surgical repair, medical treatment, or other intervention required?: | ◯ Yes<br>◯ No<br>(If unknown, please select "No". Standard procedural management (e.g. cauterization, use of prothombotic material, small vessel ligation) does not qualify as "repair.") |
| Was there a change in post-operative care due to the iAE?: | ◯ Yes<br>◯ No<br>(If unknown, please select "No") |
| Did the iAE or its management necessitate intensive care admission?: | ◯ Yes<br>◯ No<br>(If unknown, please select "No") |
| Was the intraoperative injury missed, necessitating re-operation within 7 days of index procedure?: | ◯ Yes<br>◯ No<br>(If unknown, please select "No") |
| Bleeding Related | |
| Was blood loss appreciably over normal range for procedure?: | ◯ Yes<br>◯ No<br>(If unknown, please select "No". Per Kazaryan, et al. (2013): "A normal range of blood loss for each particular procedure is subjective in a certain degree, but one can quantify it in regard to different procedures based both on contemporary scientific literature and values typical for own institution") |
| Were 2 or more units of blood products required to manage iAE?: | ◯ Yes<br>◯ No<br>(If unknown, please select "No") |
| iAE Grading | |

| | |
|---|---|
| EAUiaiC | ◯ Grade 0: Event requiring no intervention or change in operative approach, no deviation from planned intraoperative steps<br>◯ Grade 1: Events requiring change in planned intraoperative steps, not life-threatening, no tissue or organ removal. Event address in a controlled manner with no long term side effects<br>◯ Grade 2: Event requiring change in operative approach but NOT life threatening. The event was addressed in a controlled manner, however may have short/long-term side effects<br>◯ Grade 3: Event requiring deviation from planned intraoperative steps, event becoming life threatening but NOT requiring tissue or organ removal<br>◯ Grade 4: Event requiring deviation from planned intraoperative steps and with short/long-term consequences to patient<br>◯ Grade 4A: Requiring tissue or organ removal<br>◯ Grade 4B: Unable to complete planned procedure as planned due to a surgical event or technical issue or unplanned stoma<br>◯ Grade 5A: Wrong site or side open surgery or patient or no consent<br>◯ Grade 5B: Death |
| iAE Severity Classification Scheme<br><br>*Excludes minimally invasive to open conversion | ◯ Class I: Injury requiring no repair within the same procedure (e.g. cauterization, use of prothrombotic material, small vessel ligation)<br>◯ Class II: Injury requiring surgical repair, without organ removal or a change in the originally planned procedure (e.g. any suture repair, patch repair)<br>◯ Class III: Injury requiring tissue or organ removal with completion of the originally planned procedure<br>◯ Class IV: Injury requiring a significant change* and/or incompletion of the originally planned procedure<br>◯ Class V: Missed intraoperative injury requiring re-operation within 7 days<br>◯ Class IV: Intraoperative death |
| iAE Severity Classification Scheme<br><br>Suffix T<br><br>(Add if injury transfusion of ≥2 U blood) | ◯ Yes<br>◯ No |

| | |
|---|---|
| Modified Satava<br><br>†A normal range of blood loss for each particular procedure is subjective in a certain degree, but one can quantify it in regard to different procedures based both on contemporary scientific literature and values typical for own institution | ◯ Grade I: Incidents managed without change of operative approach and without further consequences for the patient. This includes minor injury of adherent or adjacent organs and minimal change of intraoperative tactics and cases with blood loss over normal range†<br>◯ Grade II: Incidents with further consequences for the patient This includes cases requiring limited resection of intraoperatively injured organs or cases with blood loss which is appreciably over normal range†. For laparoscopic/thoracoscopic/endoscopic surgery it includes intraoperative incidents requiring conversion<br>◯ Grade III: Incident leading to significant consequences for patient |
| ClassIntra ® (formerly CLASSIC)<br><br>‡Inclusive of surgery or anesthesia related events | ◯ Grade 0: No deviation from the ideal intraoperative course<br>◯ Grade I: Any deviation from the ideal intraoperative course without the need for any additional treatment or intervention. Patient asymptomatic or with mild symptoms<br>◯ Grade II: Any deviation from the ideal intraoperative course with the need for any additional minor treatment or intervention that is not life threatening and not leading to permanent disability. Patient with moderate symptoms<br>◯ Grade III: Any deviation from the ideal intraoperative course with the need for any additonal moderate or treatment or intervention which is potentially life-threatening and/or potentially leading to permanent disability. Patient with severe symptoms<br>◯ Grade IV: Any deviation from the ideal intraoperative course with the need for any additional major and urgent treatment or intervention. Patient with life threatening symptoms or leading to permanent disability<br>◯ Grade V: Any deviation from the ideal intraoperative course with death of the patient |
| EAES Classification | ◯ Grade 1: Minor error, no damage or corrective action required<br>◯ Grade 2: Minor consequential error requiring correc- tive action but no change in post-operative care<br>◯ Grade 3: Consequential error requiring major correc- tive action and/or change in post-operative pathway<br>◯ Grade 4: Life-threatening complication that requires major or immediate corrective action which led to a significant alteration of the post-operative pathway which may include re-operation or intensive care admission<br>◯ Grade 5: Major consequential error resulting in death |

**SCENARIO 10:**

**Robotic-assisted sacrocolpopexy. Post open hysterectomy**
**Complication/Management: Vaginotomy during dissection. 2-layer closure**

Patient

| | |
|---|---|
| Was the iAE associated with death of the patient?: | ◯ Yes<br>◯ No<br>(If unknown, please select "No") |
| Was the iAE immediately life-threatening?: | ◯ Yes<br>◯ No<br>(If unknown, please select "No") |
| Were there significant consequences to the patient as a result of the iAE?: | ◯ Yes<br>◯ No<br>(If unknown, please select "No". Some examples of iAEs with significant consequences includes injury to or unplanned removal of an otherwise healthy major organ (e.g. unplanned pulmonectomy, nephrectomy) and events that are exceedingly challenging to manage in a controlled manner without potential for long-term patient consequences (e.g. increased risk of postoperative multiorgan failure with major blood transfusion).) |
| Was the incorrect site, side, or surgical approach used without consent?: | ◯ Yes<br>◯ No<br>(If unknown, please select "No") |

Procedure

| | |
|---|---|
| Were there any changes in the ideal intraoperative course related to iAE?: | ◯ Yes<br>◯ No<br>(If unknown, please select "No". Changes in course include minor incidents such as unintended cauterization, equipment malfunction, unanticipated anesthesiologic challenges, or procedural delays regardless of consequence or management.) |
| Was there an unanticipated conversion of approach or significant change to the operative steps of the originally planned procedure due to iAE?: | ◯ Yes<br>◯ No<br>(If unknown, please select "No") |
| Was planned procedure aborted or incomplete due to iAE?: | ◯ Yes<br>◯ No<br>(If unknown, please select "No") |
| Unplanned stoma as a result of iAE?: | ◯ Yes<br>◯ No<br>(If unknown, please select "No") |
| Unplanned tissue or organ removal as a result of iAE?: | ◯ Yes<br>◯ No<br>(If unknown, please select "No") |

iAE Management

| | |
|---|---|
| Was any surgical repair, medical treatment, or other intervention required?: | ◯ Yes<br>◯ No<br>(If unknown, please select "No". Standard procedural management (e.g. cauterization, use of prothombotic material, small vessel ligation) does not qualify as "repair.") |
| Was there a change in post-operative care due to the iAE?: | ◯ Yes<br>◯ No<br>(If unknown, please select "No") |
| Did the iAE or its management necessitate intensive care admission?: | ◯ Yes<br>◯ No<br>(If unknown, please select "No") |
| Was the intraoperative injury missed, necessitating re-operation within 7 days of index procedure?: | ◯ Yes<br>◯ No<br>(If unknown, please select "No") |
| Bleeding Related | |
| Was blood loss appreciably over normal range for procedure?: | ◯ Yes<br>◯ No<br>(If unknown, please select "No". Per Kazaryan, et al. (2013): "A normal range of blood loss for each particular procedure is subjective in a certain degree, but one can quantify it in regard to different procedures based both on contemporary scientific literature and values typical for own institution") |
| Were 2 or more units of blood products required to manage iAE?: | ◯ Yes<br>◯ No<br>(If unknown, please select "No") |
| iAE Grading | |

| | |
|---|---|
| EAUiaiC | ◯ Grade 0: Event requiring no intervention or change in operative approach, no deviation from planned intraoperative steps<br>◯ Grade 1: Events requiring change in planned intraoperative steps, not life-threatening, no tissue or organ removal. Event address in a controlled manner with no long term side effects<br>◯ Grade 2: Event requiring change in operative approach but NOT life threatening. The event was addressed in a controlled manner, however may have short/long-term side effects<br>◯ Grade 3: Event requiring deviation from planned intraoperative steps, event becoming life threatening but NOT requiring tissue or organ removal<br>◯ Grade 4: Event requiring deviation from planned intraoperative steps and with short/long-term consequences to patient<br>◯ Grade 4A: Requiring tissue or organ removal<br>◯ Grade 4B: Unable to complete planned procedure as planned due to a surgical event or technical issue or unplanned stoma<br>◯ Grade 5A: Wrong site or side open surgery or patient or no consent<br>◯ Grade 5B: Death |
| iAE Severity Classification Scheme<br><br>*Excludes minimally invasive to open conversion | ◯ Class I: Injury requiring no repair within the same procedure (e.g. cauterization, use of prothrombotic material, small vessel ligation)<br>◯ Class II: Injury requiring surgical repair, without organ removal or a change in the originally planned procedure (e.g. any suture repair, patch repair)<br>◯ Class III: Injury requiring tissue or organ removal with completion of the originally planned procedure<br>◯ Class IV: Injury requiring a significant change* and/or incompletion of the originally planned procedure<br>◯ Class V: Missed intraoperative injury requiring re-operation within 7 days<br>◯ Class IV: Intraoperative death |
| iAE Severity Classification Scheme<br><br>Suffix T<br><br>(Add if injury transfusion of ≥2 U blood) | ◯ Yes<br>◯ No |

| | |
|---|---|
| Modified Satava<br><br>†A normal range of blood loss for each particular procedure is subjective in a certain degree, but one can quantify it in regard to different procedures based both on contemporary scientific literature and values typical for own institution | ◯ Grade I: Incidents managed without change of operative approach and without further consequences for the patient. This includes minor injury of adherent or adjacent organs and minimal change of intraoperative tactics and cases with blood loss over normal range†<br>◯ Grade II: Incidents with further consequences for the patient This includes cases requiring limited resection of intraoperatively injured organs or cases with blood loss which is appreciably over normal range†. For laparoscopic/thoracoscopic/endoscopic surgery it includes intraoperative incidents requiring conversion<br>◯ Grade III: Incident leading to significant consequences for patient |
| ClassIntra ® (formerly CLASSIC)<br><br>‡Inclusive of surgery or anesthesia related events | ◯ Grade 0: No deviation from the ideal intraoperative course<br>◯ Grade I: Any deviation from the ideal intraoperative course without the need for any additional treatment or intervention. Patient asymptomatic or with mild symptoms<br>◯ Grade II: Any deviation from the ideal intraoperative course with the need for any additional minor treatment or intervention that is not life threatening and not leading to permanent disability. Patient with moderate symptoms<br>◯ Grade III: Any deviation from the ideal intraoperative course with the need for any additonal moderate or treatment or intervention which is potentially life-threatening and/or potentially leading to permanent disability. Patient with severe symptoms<br>◯ Grade IV: Any deviation from the ideal intraoperative course with the need for any additional major and urgent treatment or intervention. Patient with life threatening symptoms or leading to permanent disability<br>◯ Grade V: Any deviation from the ideal intraoperative course with death of the patient |
| EAES Classification | ◯ Grade 1: Minor error, no damage or corrective action required<br>◯ Grade 2: Minor consequential error requiring correc- tive action but no change in post-operative care<br>◯ Grade 3: Consequential error requiring major correc- tive action and/or change in post-operative pathway<br>◯ Grade 4: Life-threatening complication that requires major or immediate corrective action which led to a significant alteration of the post-operative pathway which may include re-operation or intensive care admission<br>◯ Grade 5: Major consequential error resulting in death |

#### Collaborative authorship (optional)

| | |
|---|---|
| Would you like to submit your information for inclusion within the collaborative authorship in a future publication? | ◯ Yes<br>◯ No<br>(Note: Clicking yes will redirect you to a separate survey) |

### Collaborative Authorship Credit

Please complete the following information if you are interested in being included in as a collaborative author for the future publication of these results.

218) Last Name ____________________

219) First Name ____________________

220) Attribution ____________________
(Department, Institution, City, State, Country)

221) Email ____________________
